# Derivation and validation of a type 2 diabetes treatment selection algorithm for SGLT2-inhibitor and DPP4-inhibitor therapies based on glucose-lowering efficacy: cohort study using trial and routine clinical data

**DOI:** 10.1101/2021.11.11.21265959

**Authors:** John M Dennis, Katherine G Young, Andrew P McGovern, Bilal A Mateen, Sebastian J Vollmer, Michael D Simpson, William E Henley, Rury R Holman, Naveed Sattar, Ewan R Pearson, Andrew T Hattersley, Angus G Jones, Beverley M Shields, on behalf of the MASTERMIND consortium

**Affiliations:** University of Exeter Medical School. Address: Institute of Biomedical & Clinical Science, RILD Building, Royal Devon & Exeter Hospital, Barrack Road, Exeter EX2 5DW, UK; The Alan Turing Institute. Address: British Library, 96 Euston Road, London, NW1 2DB, UK; University College London, Institute of Health Informatics. Address: University College London, 222 Euston Rd, London NW1 2DA, London, UK; University of Warwick, Department of Statistics. Address: Department of Statistics, University of Warwick, Coventry, CV4 7AL, UK; Newcastle University, Research Software Engineering Team. Address: Newcastle University, Newcastle upon Tyne, NE1 7RU, UK; University of Exeter Medical School. Address: Health Statistics Group, Institute of Health Research, University of Exeter Medical School, Exeter EX1 2LU, UK; Diabetes Trials Unit, Oxford Centre for Diabetes, Endocrinology and Metabolism, University of Oxford. Oxford NIHR Biomedical Research Centre, Churchill Hospital, Oxford. Address: Diabetes Trials Unit, Oxford Centre for Diabetes, Endocrinology and Metabolism, University of Oxford, Oxford, UK; Institute of Cardiovascular and Medical Sciences, University of Glasgow, Glasgow G12 8TA, UK; University of Dundee. Address: Division of Molecular & Clinical Medicine, Ninewells Hospital and Medical School, University of Dundee, Dundee DD1 9SY, UK

## Abstract

**Objective:** To establish whether clinical patient characteristics routinely measured in primary care can identify people with differing short-term benefits and risks for SGLT2-inhibitor and DPP4-inhibitor therapies, and to derive and validate a treatment selection algorithm to identify the likely optimal therapy for individual patients.

**Design:** Prospective cohort study.

**Setting:** Routine clinical data from United Kingdom general practice (Clinical Practice Research Datalink [CPRD]), and individual-level clinical trial data from 14 multi-country trials of SGLT2-inhibitor and DPP4-inhibitor therapies.

**Participants:** 26,877 new users of SGLT2-inhibitor and DPP4-inhibitor therapy in CPRD over 2013-2019, and 10,414 participants randomised to SGLT2-inhibitor or DPP4-inhibitor therapy in 14 clinical trials, including 3 head-to-head trials of the two therapies (n=2,499).

**Main outcome measures:** The primary outcome was achieved HbA1c 6 months after initiating therapy. Clinical features associated with differential HbA1c outcomes with SGLT2-inhibitor and DPP4-inhibitor therapies were identified in routine clinical data, with associations then tested in trial data. A multivariable treatment selection algorithm to predict differential HbA1c outcomes was developed in a CPRD derivation cohort (n=14,069), with validation in a CPRD validation cohort (n=9,376) and the head-to-head trials. In CPRD, we further explored the relationship between model predictions and secondary outcomes of weight loss and treatment discontinuation.

**Results:** The final treatment selection algorithm included HbA1c, eGFR, ALT, age, and BMI, which were identified as predictors of differential HbA1c outcomes with SGLT2-inhibitor and DPP4-inhibitor therapies using both routine and trial data. In validation cohorts, patient strata predicted to have a ≥5 mmol/mol HbA1c reduction with SGLT2-inhibitor therapy compared with DPP4-inhibitor therapy (38.8% of CPRD validation sample) had an observed greater reduction of 8.8 mmol/mol [95%CI 7.8-9.8] in the CPRD validation sample, a 5.8 mmol/mol (95%CI 3.9-7.7) greater reduction in the Cantata D/D2 trials, and a 6.6 mmol/mol [95%CI 2.2-11.0]) greater reduction in the BI1245.20 trial. In CPRD, there was a greater weight reduction with SGLT2-inhibitor therapy regardless of predicted glycaemic benefit. Strata predicted to have greater reduction in HbA1c on SGLT2-inhibitor therapy had a similar risk of discontinuation as on DPP4-inhibitor therapy. In contrast, strata predicted to have greater reduction in HbA1c with DPP4-inhibitor therapy were half as likely to discontinue DPP4-inhibitor therapy than SGLT2-inhibitor therapy.

**Conclusions:** Routinely measured clinical features are robustly associated with differential glycaemic responses to SGLT2-inhibitor and DPP4-inhibitor therapies. Combining features into a treatment selection algorithm can inform clinical decisions concerning optimal type 2 diabetes treatment choices.

**Key messages:** *What is already known on this subject:*

- Despite there being multiple glucose-lowering treatment options available for people with type 2 diabetes, current guidelines do not provide clear advice on selecting the optimal treatment for most patients.
- It is unknown whether routinely measured clinical features modify the risks and benefits of two common treatment options, DPP4-inhibitor or SGLT2-inhibitor therapy, and which could be used to target these treatments to those patients most likely to benefit.

*What this study adds:*

- Using data from 10,414 participants in 14 randomised trials, and 26,877 patients in UK primary care, we show several routinely available clinical features, notably glycated haemoglobin (HbA1c) and kidney function, are robustly associated with differential HbA1c responses to initiating SGLT2-inhibitor and DPP4-inhibitor therapies.
- Combining clinical features into a multivariable treatment selection model identifies validated patient strata with 1) a >5 mmol/mol HbA1c benefit for SGLT2-i therapy compared with DPP4-inhibitor therapy ; 2) a 50% reduced risk of early treatment discontinuation with DPP4-inhibitor therapy compared with SGLT2-inhibitor therapy.
- Our findings demonstrate a precision medicine approach based on routine clinical features can inform clinical decisions concerning optimal type 2 diabetes treatment choices.

## Introduction

SGLT2-inhibitors (SGLT2i) and DPP4-inhibitors (DPP4i) are recommended glucose-lowering treatment options after metformin in all major type 2 diabetes clinical guidelines.(1, 2) They represent around 60% of second-line treatment initiations in the UK,(3) and 27% in the US,(4). Trial data suggest that the average glucose-lowering efficacy of both therapies is similar, although SGLT2i are associated with weight loss, whilst DPP4i may be weight neutral.(5) Differences in tolerability have not been evaluated in large numbers of patients in routine practice. Whilst current ADA/EASD guidelines recommend SGLT2i and/or GLP-1 receptor agonists in people with established atherosclerotic cardiovascular disease, heart failure, or chronic kidney disease,(1) this stratification only applies to up to 15–20% of people with type 2 diabetes.(6, 7) This means that, for a majority of people with type 2 diabetes, there is considerable uncertainty on optimal treatment after initial metformin.

A potential approach to treatment selection in type 2 diabetes is to use individual-level patient characteristics to target specific glucose-lowering treatment to those people most likely to benefit.(8) The hope is that such a targeted ‘precision medicine’ approach will, when implemented, improve glucose-lowering efficacy and drug tolerability, reduce side-effects, and reduce the risk of developing diabetes complications.(9) Recent studies have shown the potential for type 2 diabetes precision medicine, identifying subgroups with different glycaemic response to specific agents, rates of glycaemic progression, and risk of complications.(10-12) However, previous studies have not evaluated the clinical utility of the proposed precision medicine approaches, in particular whether differential outcomes between drug classes can be robustly predicted.(13)

In this study we aimed to build on recently proposed ‘effect-modelling’ approaches to detect patient-level treatment effects(14-16) to evaluate: 1) whether individual routine clinical features are robustly associated with differential glycaemic response to SGLT2i and DPP4i therapies; 2) whether combining clinical features to predict HbA1c responses to SGLT2i and DPP4i therapies can inform selection of treatment based on optimal glucose-lowering; 3) whether selecting treatments based on predicted HbA1c responses relates to changes in weight and the likelihood of treatment discontinuation.

## Methods

### Study population

#### Routine clinical data

Patients initiating SGLT2i and DPP4i therapies after January 1^st^, 2013, were identified in UK Clinical Practice Research Datalink (CPRD) GOLD (July 2019 download),(17) following our published protocol.(18) We then excluded those prescribed a SGLT2i or DPP4i as first-line treatment (as not recommended in UK guidelines)(2), co-treated with insulin, with eGFR <45 (where use of SGLT2i is usually contraindicated), or without a valid baseline HbA1c (<53 or ≥ 120 mmol/mol) value (**sFlowchart**). The following baseline clinical features were extracted, chosen due to availability in the majority of patients: HbA1c (most recent value in previous 6 months or 7 days after treatment initiation), age at treatment initiation, sex, diabetes duration, BMI, weight, c, eGFR, HDL-cholesterol, triglycerides, ALT, albumin, and bilirubin (all most recent value in the 2 years prior to therapy initiation). We also identified the number of currently prescribed glucose-lowering treatments, and the number of glucose-lowering drug classes ever prescribed.

#### Clinical trial data

Individual participant data from 14 randomised trials of SGLT2i and DPP4i therapies (total participant n=10,414) were accessed from the Yale University Open Data Access Project and Vivli.(19, 20) This included three active comparator HbA1c efficacy trials of SGLT2i vs DPP4i treatment (CANTATA-D and CANTATA-D2 trials of canagliflozin [SGLT2i] versus sitagliptin [DPP4i] /placebo; placebo arms not analysed),(21, 22) the BI1245.20 trial of empagliflozin [SGLT2i] versus sitagliptin),(23) six efficacy trials of SGLT2i versus placebo/sulfonylurea (four canagliflozin trials,(24-27) two empagliflozin trials;(28, 29) non-SGLT2i treatment arms not analysed), the EMPA-REG OUTCOME cardiovascular outcome trial (empaglifozin versus placebo; placebo arm not analysed, patients with insulin co-treatment excluded, and only HbA1c measures on unchanged glucose-lowering therapy analysed),(30) and four efficacy trials of DPP4i versus placebo/sulfonylurea (all linagliptin, non-DPP4i treatment arms not analysed).(31-34) Participants randomised to different doses of active agents were pooled for analysis. Full details of trial inclusion criteria and final study cohorts are provided in **sTable 1** and **Supplement 2**. HbA1c outcome data, and baseline assessment data for the same clinical features as in CPRD, were extracted.

### Outcomes

The primary efficacy outcome was the HbA1c value achieved six months after drug initiation, adjusted for baseline HbA1c.(35) In CPRD, this outcome was defined as the closest HbA1c to six months (within 3-15 months to maximise the number of patients with a valid outcome) after initiation, on unchanged therapy. In clinical trials, this outcome was defined as all on treatment HbA1c values at study visits from 3-6 months after randomisation.

Secondary outcomes in CPRD comprised: 1) achieved weight six months after initiation (closest recorded weight to six months (within 3-15 months); 2) treatment discontinuation within six months of drug initiation (a proxy of drug tolerability). Patients were required to have three months of follow-up time after their last prescription to confirm the drug was discontinued.

### Statistical analysis

#### Identifying individual features associated with differential drug response

In CPRD, linear regression models were used to estimate the association between baseline HbA1c and six-month HbA1c by drug, fitting a drug-by-baseline HbA1c interaction term, and baseline HbA1c modelled as a 3-knot restricted cubic spline (RCS) to allow for non-linearity. In this model and all subsequent CPRD models, we controlled for differences in drug order and potential adherence effects by adjusting for the number of current glucose-lowering medications, and the number of previously initiated medications. We also adjusted for differences in month of outcome measurement by adjusting for the month (relative to baseline) that the HbA1c outcome was recorded. We then sequentially assessed associations by drug for other baseline clinical features, by adding each in-turn as drug-by-feature interactions to the baseline HbA1c adjusted model. Each feature was standardised to allow comparison of effect size across features. We conducted a complete case analysis for each feature of interest. To evaluate model fit we visually examined normality of residuals and linearity of associations for continuous variables.

In the trials, we estimated associations for the same clinical features as per CPRD using repeated measures mixed effect models and patient-level random-effects, using on-therapy HbA1c values at each discrete study visit up to six months post-randomisation. Features were standardised to CPRD distributions. For active comparator trials, we compared efficacy by drug using drug-by-feature interaction terms. For each clinical feature, trial estimates were then pooled using two-stage random effects meta-analysis.(36)

#### Treatment selection model derivation

We set out to develop a counterfactual prediction model to predict HbA1c outcome for an individual patient if they were to receive either SGLT2i or DPP4i therapy (treatment selection model). A multivariable linear regression model was developed in a CPRD 60% random sample (derivation cohort). The remaining 40% of patients were held back for evaluating model performance. By including treatment-by-feature interaction terms, the model facilitated prediction of the outcome on each treatment, conditional on the features included as interaction terms, and thus enabled prediction of individualised treatment effects. For each person, the difference between the predicted HbA1c outcome on each therapy provides an estimate of the ‘individualised’ treatment effect.

To inform variable selection, an initial linear regression model for six-month HbA1c with interaction terms between treatment and all baseline clinical features (continuous features modelled as three-knot RCS) as explanatory variables was fitted. The relative importance of each baseline clinical feature for estimating individualised treatment effects was assessed by estimating the proportion of chi-squared explained by the interaction term for each feature (which represents the differential effect of the feature on HbA1c outcome), with bootstrapped confidence intervals. Stepwise forward selection was used to identify the feature subset for the final model, by adding drug-by-feature interaction terms in order of relative importance to a base model that included all clinical features without interaction terms, with inclusion based on a p<0.01 threshold. Non-differential features were omitted from the final model as they explained little variation in predicting overall HbA1c outcome (additional R^2^ 0.004). To adjust for overfitting in the final model, penalised ridge regression was used to optimise for AIC.(37) A complete case approach was used throughout as missing baseline data were considered likely to be missing not at random.(38) Standard performance metrics were estimated to assess model performance for predicting HbA1c outcome; optimism-adjusted model R^2^, root mean square error, and the calibration slope and calibration-in-the-large.(39)

#### Treatment selection model external validation

In contrast to standard prediction models, for treatment selection models, accurately predicting the magnitude of difference between therapies (the individualised treatment effect) is more important than accurately predicting the outcome.(40) Standard model performance metrics test the ability of a model to predict the outcome, and are therefore of limited use in the context of evaluating a model estimating individualised treatment effects.(40, 41) The challenge for validation is that, without a crossover trial, the difference in treatment effect cannot be measured directly within an individual, as each individual will have outcome data available on the treatment initiated, but not on the other treatment(s) they could have but did not initiate (the counterfactual outcome).

What is possible is evaluation of a treatment selection model in validation data by assessing differences in observed outcome in patient strata defined by model-predicted individualised treatment effects. Evaluation using this approach for the SGLT2i versus DPP4i treatment selection model is described in **Figure 1**, and was applied to evaluate the model in individual-level independent validation data from three randomised SGLT2i versus DPP4i head-to-head trials, and the CPRD 40% hold-back set. CANTATA-D and CANTATA-D2 trial participants were pooled (these differed only in background therapy [D: metformin only; D2: metformin and sulfonylurea]); BI1245.20 participants (drug-naïve to glucose-lowering agents) were analysed separately. The following steps were undertaken in each dataset: (1) HbA1c reductions were predicted for both SGLT2i and DPP4i therapies for all individuals; (2) predictions were used to estimate individualised treatment effects (the estimated difference in HbA1c outcome on the two therapies); (3) strata were defined by decile of predicted individualised treatment effect, and by clinically defined HbA1c cut-offs of predicted individualised treatment effect (SGLT2i benefit: ≥10, 5-10, 3-5, 0-3 mmol/mol; DPP4i benefit: ≥5, 3-5, 0-3 mmol/mol); 4) linear regression models were used to contrast within-strata HbA1c outcome in concordant (i.e. therapy received is the therapy predicted to have greatest HbA1c lowering) versus discordant (i.e. therapy received is the predicted non-optimal therapy) subgroups. In CPRD, estimates were adjusted for clinical features in the treatment selection model (to improve precision and control for potential differences in covariate balance within subgroups). Trial estimates were unadjusted. In the randomised trials, the outcome assessed was last-observational-carried-forward six-month HbA1c. Longer term 12-month HbA1c outcome was evaluated in the trials as a sensitivity analysis.

**Figure 1:**
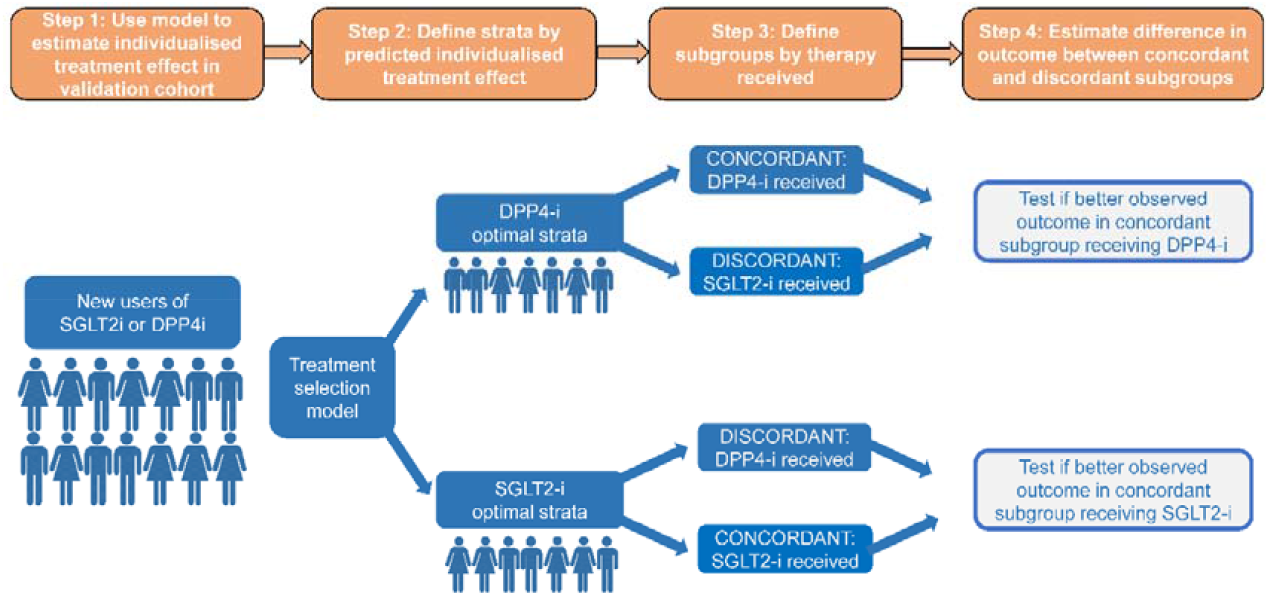
SGLT2-inhibitor versus DPP4-inhibitor treatment selection model evaluation framework. In a suitable validation dataset (Step 1), model derived individualised treatment effects can be used to assign patients into strata based on model-predicted optimal therapy (Step 2). In its simplest form each patient can be assigned one of to two strata: A) DPP4i is the predicted optimal therapy; B) SGLT2i is the predicted optimal therapy. Concordant (therapy received = predicted optimal therapy) and discordant (therapy received ≠ predicted optimal therapy) subgroups can then be defined based on the therapy actually received by each patient (Step 3). Finally, to evaluate treatment selection model performance, the magnitude of improvement in outcome in the concordant compared to the discordant subgroups can be estimated within each strata (Step 4). More granular strata can be defined based on the size of predicted treatment effect (for example, a strata defined by a predicted >5 mmol/mol HbA1c benefit on SGLT2i versus DPP4i), and the same concordant-discordant framework can be used to evaluate if the within-strata observed difference in outcome is similar to the predicted difference.

#### Weight change and treatment discontinuation

In CPRD, to assess whether selecting treatment based on predicted HbA1c outcome altered other short-term patient outcomes, we tested whether six-month weight change and risk of discontinuation varied by degree of predicted individualised HbA1c treatment effect. The same concordant-discordant approach as for HbA1c outcome was used. For treatment discontinuation, a logistic regression model was fitted with predicted HbA1c difference between therapies as the exposure (3-knot RCS), adjusting for baseline weight, the number of current glucose-lowering medications, and the number of previously initiated medications. For weight change, a linear regression was fitted with additional adjustment for baseline weight. Longer term 12-month outcomes were evaluated as a sensitivity analysis.

All analyses were conducted using Rv4.0.2. CPRD data preparation was carried out using Stata v15.0. We followed TRIPOD (transparent reporting of a multivariable model for individual prognosis or diagnosis) guidance for model development and reporting.(39)

### Patient and public involvement

People with type 2 diabetes were involved in the MASTERMIND consortium and were key in identifying that better, more tailored, evidence was needed for the choice of second and third-line glucose-lowering therapy. There was no patient or public involvement when conducting this specific study in terms of study design, analysis, interpretation or writing.

## Results

### Glycaemic response: Baseline HbA1c, current age, diabetes duration, eGFR, ALT and BMI are individual clinical features associated with differential HbA1c response with SGLT2i and DPP4i therapy

Baseline clinical characteristics are reported in **sTable 1** for all study cohorts. The final CPRD cohort comprised 10,253 new users of SGLT2i therapy and 16,624 new users of DPP4i therapy with valid HbA1c outcome data (**sFlowchart**). In CPRD, higher baseline HbA1c was associated with a markedly greater HbA1c reduction at 6 months with SGLT2i compared to DPP4i (**Figure 2**). This association was replicated in the active comparator SGLT2i versus DPP4i trials (**Figure 2**). Adjusted for baseline HbA1c, multiple individual features showed evidence of differential responses in CPRD (**Figure 3a**). Differential effects of greatest magnitude were seen for current age and diabetes duration (higher age / longer duration associated with greater response to DPP4i but not SGLT2i), eGFR and ALT (higher values associated with greater response to SGLT2i, lesser response to DPP4i), and BMI (higher BMI associated with a lesser response to DPP4i, no association with SGLT2i response). Differential treatment effects for BMI and ALT were replicated in trials, as was the association between higher eGFR and greater SGLT2i response (**Figure 3b**). Associations between higher age and lower eGFR with greater DPP4i response was not replicated (**Figure 3b**).

**Figure 2:**
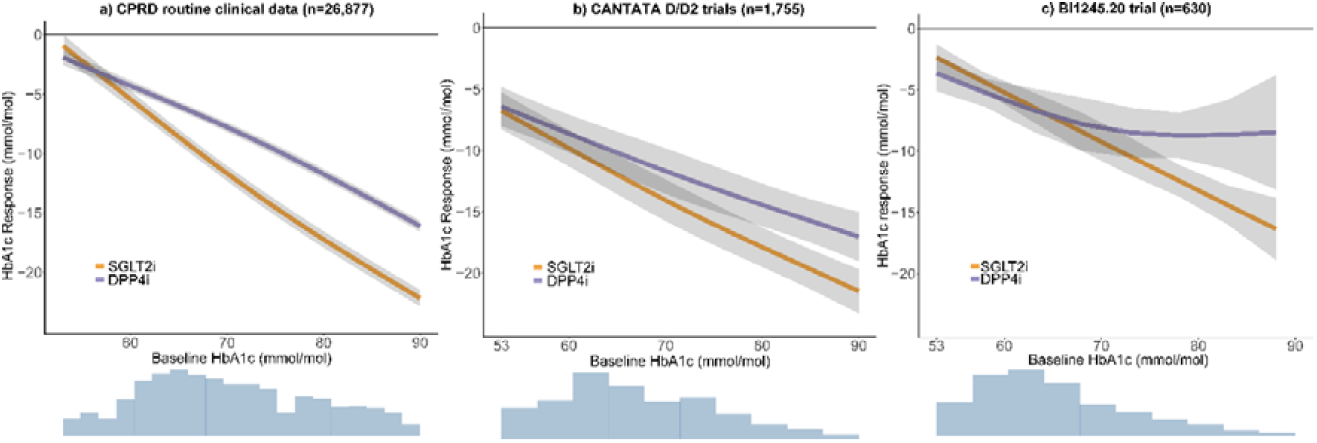
Association between baseline HbA1c and 6 month HbA1c response (outcome HbA1c – baseline HbA1c) for SGLT2-inhibitor and DPP4-inhibitor treatment a) CPRD routine clinical data (n=26,877); b) CANTATA D/D2 trials (n=1,755); c) BI1245.20 trial (n=630). Histograms show the distribution of baseline HbA1c in each dataset.

**Figure 3:**
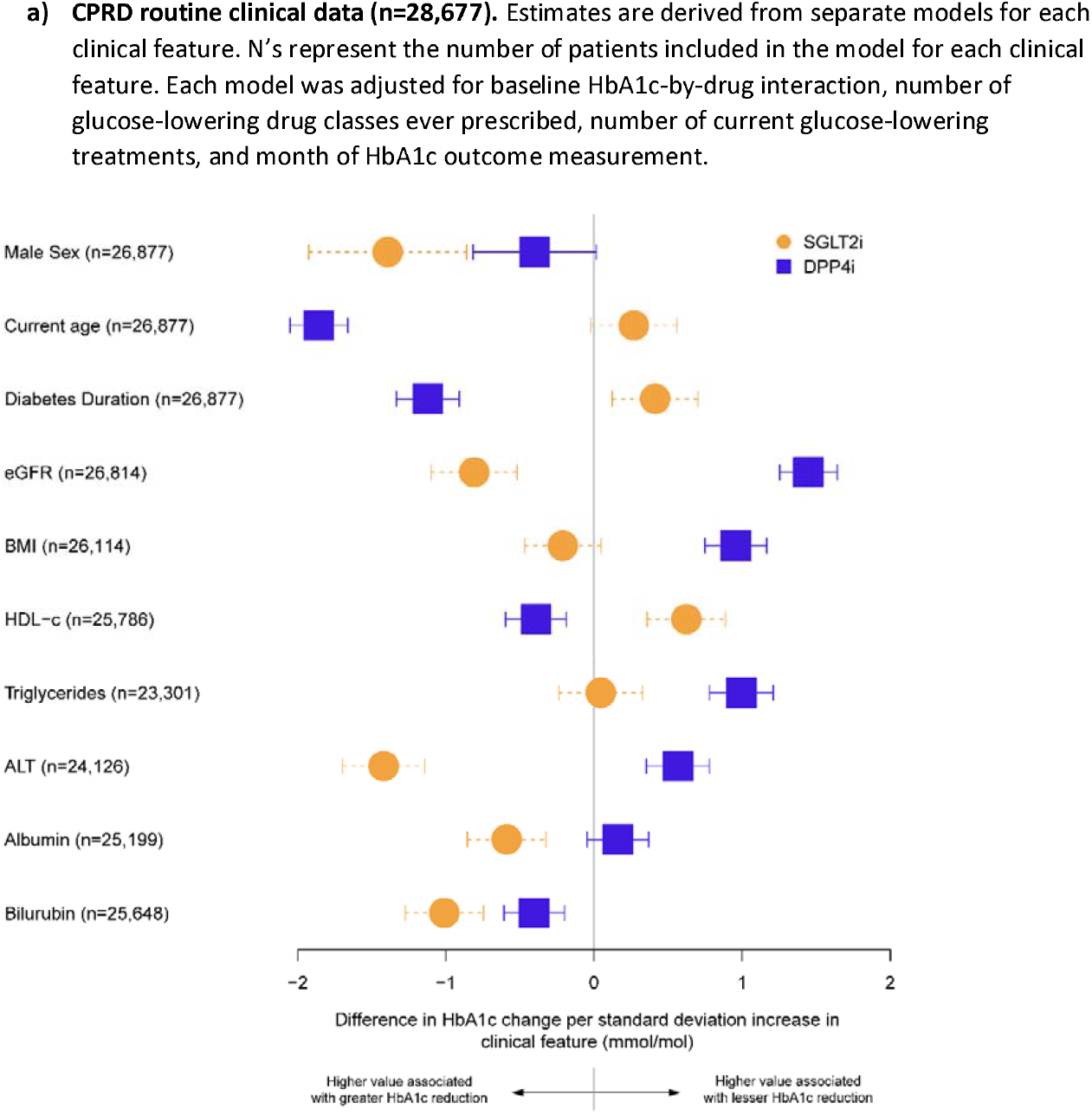
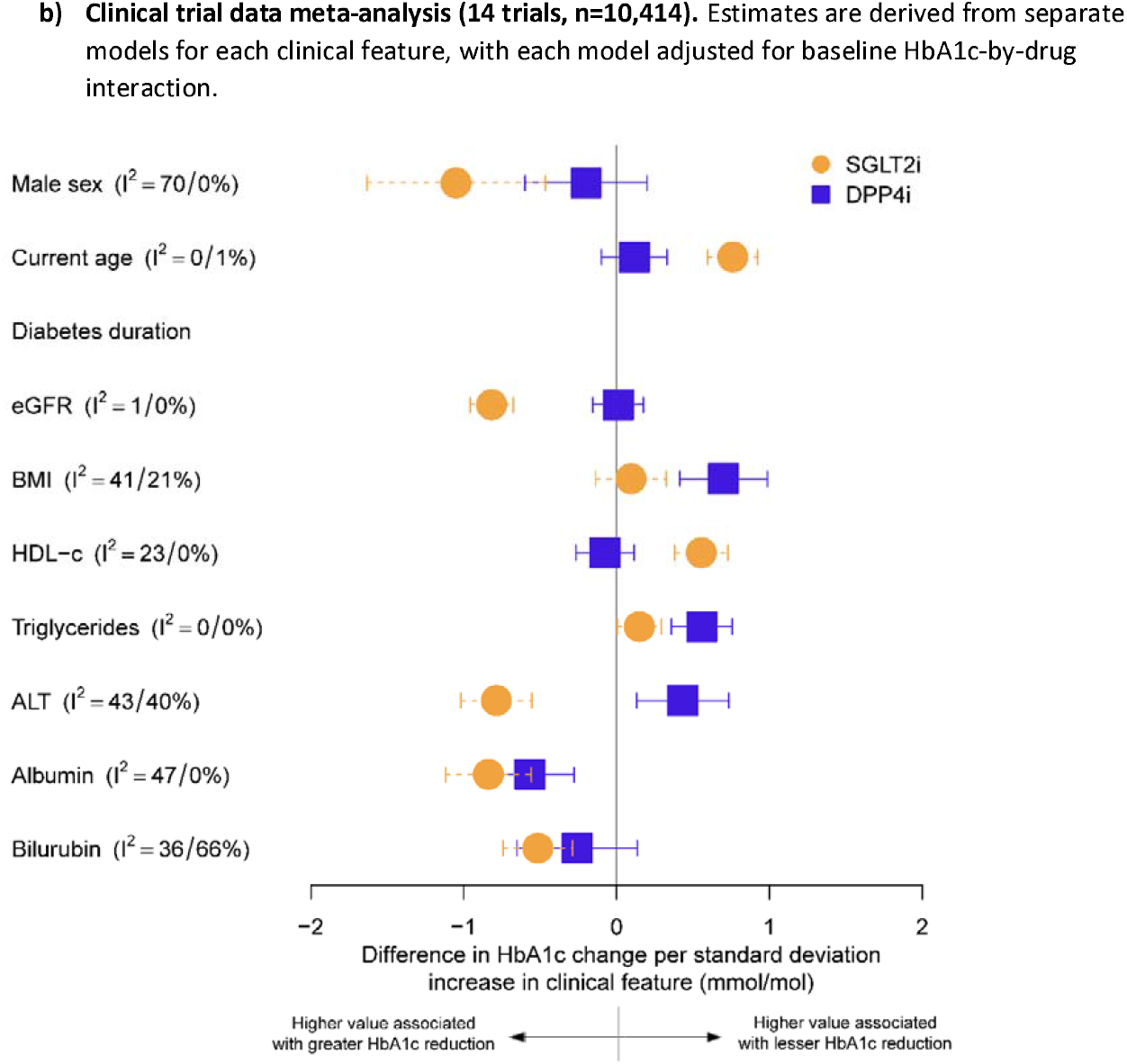
Associations between clinical features and baseline HbA1c adjusted 6 month HbA1c response for SGLT2-inhibitor and DPP4-inhibitor treatment. Negative estimates represent an association between a greater value of the clinical feature and greater HbA1c improvement, positive estimates represent an association between a greater value of the clinical feature and lesser HbA1c improvement. Data underlying the plot, including estimates from each individual trial, are reported in **sTable 2**.

#### Model development

In the model assessing all clinical features, baseline HbA1c and eGFR were the most important clinical features for predicting differential treatment effects, followed by ALT, BMI, and current age (**sFigure 1**). Only these five differential features significantly improved prediction of HbA1c outcome and were included in the final model (derivation cohort n=14,069). The full model equation is reported in **sTable 3**, with non-linear associations for continuous clinical features reported in **sFigure 2**. Performance of the model for predicting HbA1c outcome in the derivation cohort is reported in **sTable 4a**. Internal validation showed that the final model explained 29.1% of the variation in HbA1c outcome, and was well calibrated (slope 0.9967 [1=perfect]) (**sFigure 3a**).

There was evidence of marked heterogeneity in predicted individualised treatment effects, with the model predicting a benefit with SGLT2i for 84% (n=11,814) and a benefit with DPP4i for 16% (n=2,255) of patients in the derivation cohort (**Figure 4**). Across deciles of predicted individualised treatment effect, there was good calibration between observed HbA1c differences and predictions (**Figure 4**).

**Figure 4:**
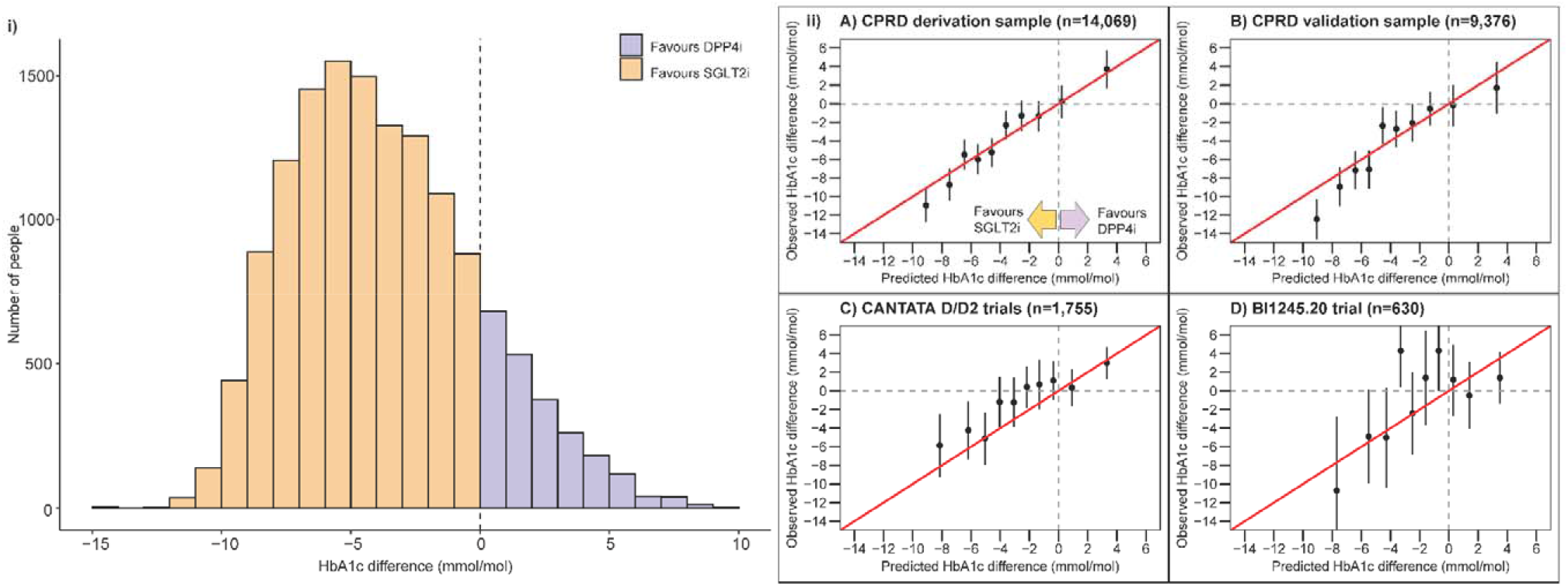
Final treatment selection model performance. **i) Distribution of the predicted individualised treatment effect of SGLT2-inhibitor treatment compared to DPP4-inhibitor treatment in the CPRD derivation sample (n=14,069) [left panel]**. Negative values reflect a predicted glucose-lowering treatment benefit on SGLT2-inhibitor treatment, positive values reflect a predicted treatment benefit on DPP4-inhibitor treatment. **ii) Calibration between observed and predicted treatment effects, by decile of predicted treatment effect [right panel], in a) CPRD derivation sample (n=14,069); b) CPRD validation sample (n=9,276); c) CANTATA D/D2 trials (n=1,755); d) BI1245.20 trial (n=630)**. In CPRD, estimates are adjusted for clinical features in the treatment selection model (to improve precision and control for potential differences in covariate balance within subgroups). Trial estimates are unadjusted.

#### Model validation: Combined clinical features have clear utility for predicting individualised HbA1c treatment effects

Calibration between observed HbA1c differences and predictions was good in CANTATA-D/D2 trials and the CPRD validation sample, in the smaller BI1245.20 trial overall calibration was less good but patient deciles with the greatest predicted HbA1c benefit on SGLT2i had a clear observed benefit in-line with predictions **(Figure 4)**. In CPRD, SGLT2i-optimal concordant patients (those who received SGLT2i and for whom SGLT2i was the predicted best therapy) had a 5.4 mmol/mol benefit compared to SGLT2i-optimal discordant patients (those who received DPP4i but for whom SGLT2i was the best predicted therapy) **(Figure 5)**. SGLT2i-optimal patients with a predicted SGLT2i benefit ≥5 mmol/mol (n=3,756 [40.1% of total]) had an 8.9 mmol/mol observed benefit if they received SGLT2i compared to DPP4i.

**Figure 5:**
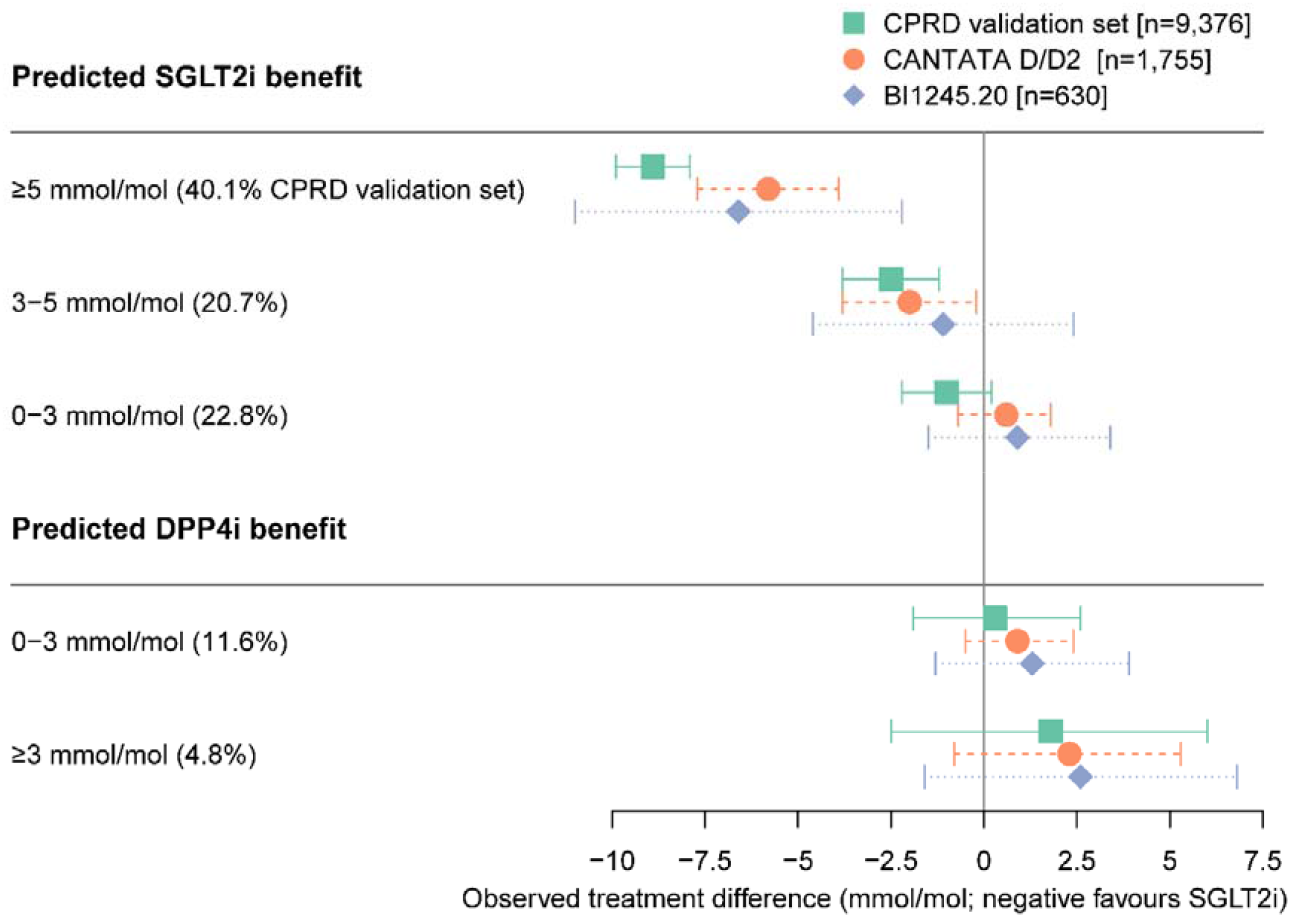
Observed treatment effects across subgroups defined by clinical cut-offs of predicted treatment benefit. In CPRD, estimates are adjusted for clinical features in the treatment selection model, trial estimates are unadjusted. **sTable 6** reports the data underlying the plot. **sFigure 4** reports the full distribution of predicted treatment difference estimates.

The model also identified a smaller group of patients with a potential HbA1c benefit on DPP4i therapy. In the CPRD validation set, DPP4i-optimal concordant patients with any predicted benefit on DPP4i did not have an observed benefit (0.6 [95%CI -1.4, 2.3] mmol/mol, n=1,540 [16.4%]) **(Figure 5)**. Those with a predicted benefit ≥3 mmol/mol on DPP4i (n=450 [4.8%]) had a 1.8 [95%CI - 2.5, 6.0] mmol/mol observed benefit. Results for both SGLT2i and DPP4i-optimal groups were similar in both trials, for 6 month **(Figure 5)** and 12-month HbA1c outcome **(sTable 6)**.

Model performance for predicting HbA1c outcome in all validation sets is reported in **sTable 4b** and **sFigure 3b-d**; calibration was good in the CPRD validation set but observed HbA1c outcome was consistently lower (better) than predicted HbA1c outcome in the trials, potentially reflecting greater adherence in trial participants.

#### Weight change does not vary by predicted individualised HbA1c treatment effect, but patients for whom DPP4i is the predicted optimal treatment based on HbA1c have a lower risk of discontinuation on DPP4i than SGLT2i therapy

In CPRD overall, at 6 months patients initiating SGLT2i had a greater median weight loss (3.7kg [IQR 3.2-4.3]) than patients initiating DPP4i (1.0kg [IQR 0.5-1.6) **(sTable 7a)**. There was greater weight loss with SGLT2i then DPP4i across all subgroups defined by model predicted differences between therapies in HbA1c outcome **(Figure 6a)**. Overall treatment discontinuation within 6 months was similar on SGLT2i and DPP4i (median 16.1% [IQR 13.5-20.3] and 14.4% [IQR 12.9-16.7] respectively), and in patients predicted to have an HbA1c benefit with SGLT2i over DPP4i (median 15.2% [IQR 13.2-20.3] and 14.4% [IQR 12.9-16.7] respectively) **(Figure 6b, sTable 7b)**. In patients predicted to have an HbA1c benefit on DPP4i, median discontinuation was lower on DPP4i than SGLT2i (14.8% [IQR 12.9, 16.8] and 26.8% [IQR 23.4-31.0] respectively). In patients with a predicted HbA1c benefit of ≥3 mmol/mol on DPP4i, discontinuation on DPP4i was half of that observed for SGLT2i (DPP4i 14.9% [IQR 13.0-16.9] versus SGLT2i 33.1% [IQR 29.7-36.9]) **(Figure 6b, sTable 7b)**. Differences were consistent at 12 months **(sFigure 5)**.

**Figure 6:**
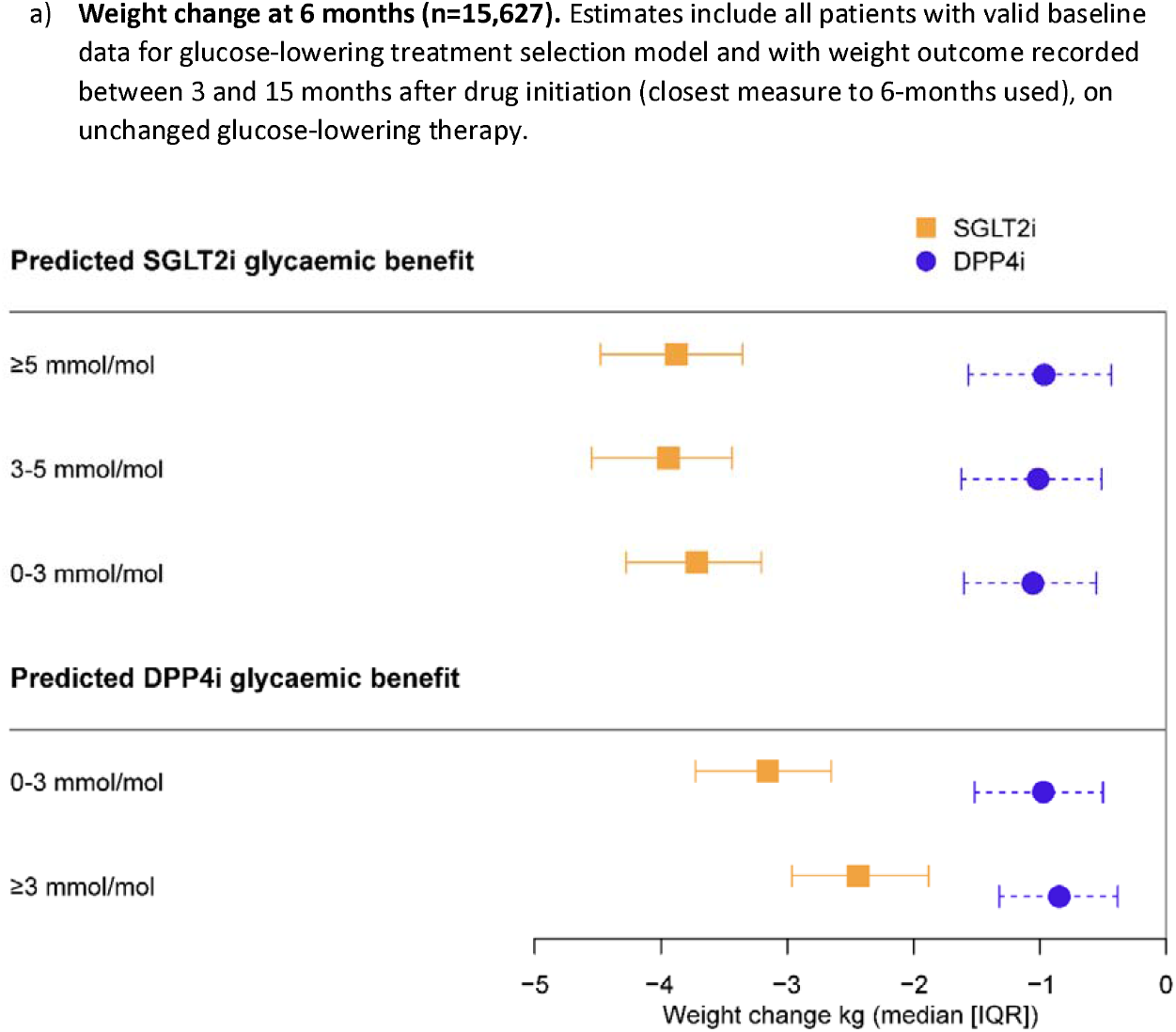
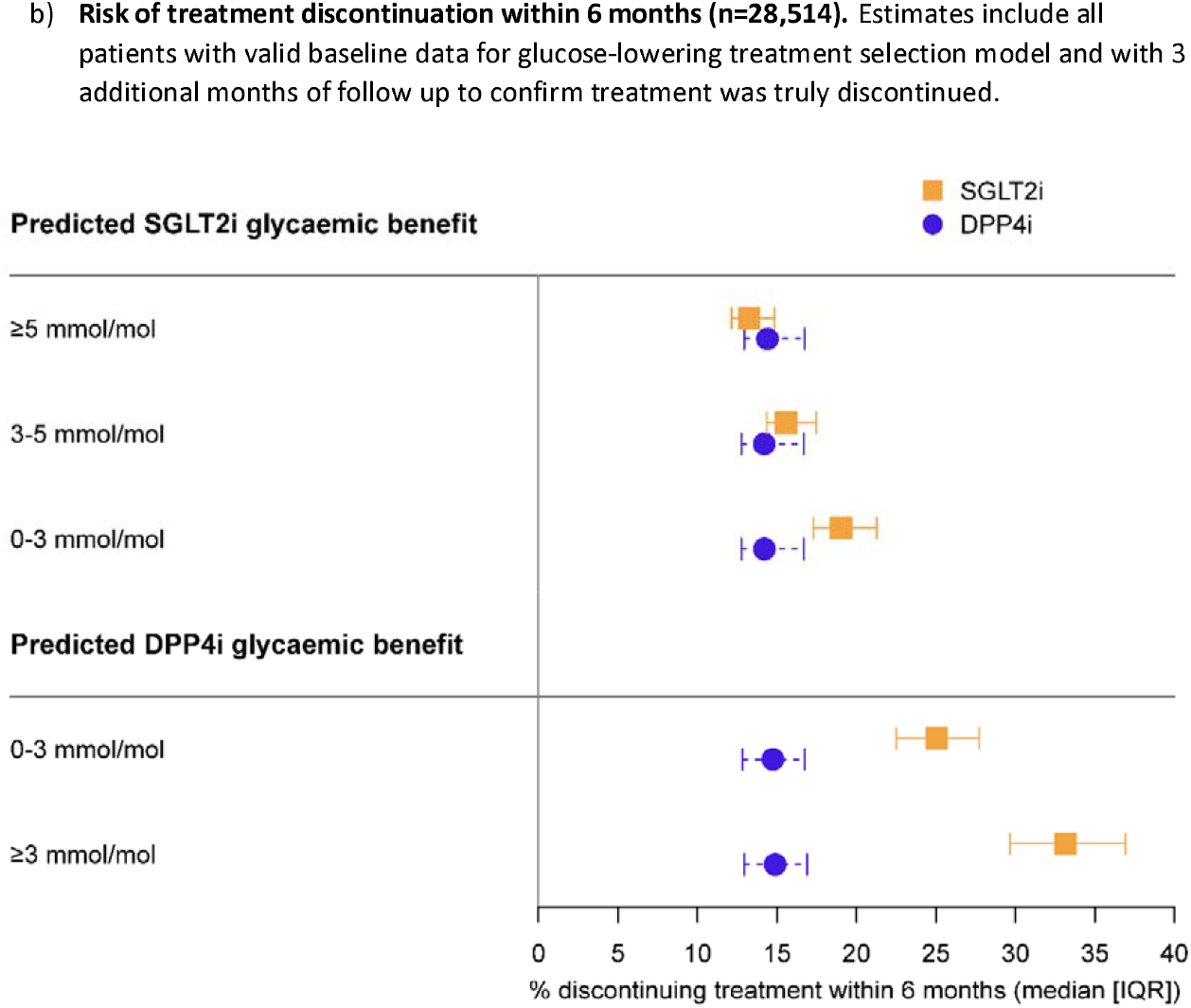
6 month weight change and risk of treatment discontinuation, across subgroups defined by clinical cut-offs of predicted treatment benefit, in CPRD routine clinical data. Data are median (interquartile range) for each subgroup. 12 month outcomes are reported in **sFigure 4**.

#### Clinical characteristics by subgroups defined by model predicted HBA1c differences between therapies

In the overall CPRD cohort (n=36,454 with valid data to fit the treatment selection model; **sTable 8**), patients with a predicted HbA1c benefit ≥5 mmol/mol with SGLT2i than DPP4i therapy (40.8% of all patients) were younger (median age 55), predominantly male (66.6%) and obese (median BMI 34.1), with higher HbA1c (median 80.2 mmol/mol), eGFR (median 97) and ALT (median 37.0 IU/L) levels **(sTable 8)**. Conversely, patients with a predicted HbA1c benefit ≥3 mmol/mol with DPP4i than SGLT2i therapy (5.0% of all patients) were older (median age 79), equally male and female (51.5% male), slimmer (median BMI 26.5), with lower HbA1c (median 61.0), eGFR (median 59) and ALT median (15.0). 50.2% of patients with a predicted HbA1c benefit ≥5 mmol/mol with SGLT2i received a SGLT2i, whilst 88.7% of patients with a predicted HbA1c benefit ≥3 mmol/mol with DPP4i received a DPP4i.

#### Development of prototype treatment selection decision aid

A research-only web tool implementing the algorithm developed for this study is available at: diabetes-calculator.uksouth.cloudapp.azure.com/calculator/

## Discussion

This study, using UK clinical data with confirmation of findings in multiple clinical trial datasets, demonstrates that routinely measured clinical features of people with type 2 diabetes are associated with differential HbA1c responses to SGLT2i and DPP4i therapies. By developing an algorithm combining baseline HbA1c, current age, BMI, eGFR and ALT, we identify a large group of people (40.8% of UK patients initiating these therapies) with a predicted glycaemic benefit ≥5 mmol/mol and greater weight loss on SGLT2i compared with DPP4i, and a similar risk of early discontinuation for both agents. We also identify a smaller group of patients (16.5% of UK patients, so around 1 in 6) with a 50% lower risk of short-term discontinuation (around 10% lower) on DPP4i compared with SGLT2i, and who may have a greater glycaemic reduction with DPP4i therapy but greater weight loss with SGLT2i. The remaining patients have a similar glycaemic response and similar risk of treatment discontinuation with both therapies, but with greater weight loss on SGLT2i.

Our analysis provides a demonstration of translational precision medicine in informing the selection of add on type 2 diabetes therapies. Validation of findings in randomised clinical trial data and an independent sample of routine patient data provides a robust demonstration of the clinical utility of using simple clinical features to provide individualised estimates of the risks and benefits of SGLT2i and DPP4i treatments.. Although not all differences in effects for individual clinical features observed in routine data were replicated in the trials, notably associations between higher current age and lower eGFR with lesser DPP4i response, the multivariable algorithm performed well in validation. The validation framework applied is analogous to cardiovascular risk prediction models such as QRISK which are used routinely in UK current practice,(42) where effectiveness reflects the ability of the model to accurately quantify risk at a population level, rather than precisely define the time to cardiovascular event for an individual. For deployment, the use of only routinely measured patient-level characteristics has large advantages in cost and feasibility compared to approaches based on non-routine measurements such as genetic information. Of critical importance, the individual-level estimates provided by the algorithm are not intended to be prescriptive, but instead to support more informed discussion on the benefits and risk of SGLT2i and DPP4i treatment for an individual patient, alongside understanding of average-level class effects, in particular the demonstrated cardiovascular benefit of SGLT2i in people with, or at high-risk of, cardiovascular disease.

The combination of clinical features predicting differential glycaemic response most likely relates to differences in the underlying mechanism of action of the two therapies, although this needs further study. For example, increased urinary glucose excretion offers a likely explanation for the greater response to SGLT2i in patients with higher levels of baseline HbA1c and kidney function, and it is biologically plausible that factors associated with insulin resistance (i.e. BMI, ALT levels) would reduce response to an agent that acts primarily through potentiating insulin secretion (DPP4i) but not SGLT2i which acts though an independent mechanism.(43)

Whilst the two-step approach of model development and validation in independent datasets used in this study is similar to that proposed in the recent Predictive Approaches to Treatment effect Heterogeneity (PATH) statement,(15) our design differs in that we used routine clinical data rather than clinical trial data for the initial ‘discovery’ analysis and model development. The advantage of using routine data for detecting and estimating heterogenous treatment effects was the large sample size and greater heterogeneity of patients in clinical practice compared to trial settings. The performance of the algorithm developed was then tested in multiple randomised trial datasets where systematic participant follow-up is available, and likelihood of confounding is much lower. Whilst our study highlights a potential for using routine health data for exploratory studies of treatment effect heterogeneity, further research on optimal designs for treatment effect heterogeneity studies is needed,(15) and given the known biases of routine data, the second step of validation in randomised data will likely always be required to demonstrate clinical utility.

Our study has several limitations. Notably, we were not able to validate differential treatment effects at the individual level. This is because individual level treatment effects are not directly observable as patient outcomes can only be observed for the treatment taken, and not the counterfactual outcome on other treatment(s) that could have been taken (the fundamental problem of causal inference).(44) Randomised crossover-trial designs are the only way to evaluate such individual level effects. In the absence of such trials, the demonstration of model utility in multiple independent datasets we provide is the next best evidence. We did not evaluate differential treatment effects by ethnicity, due to the limited numbers of non-white patients in both the routine clinical and trial datasets, an important area for future work. We also did not evaluate longer-term patient outcomes, in particular cardiovascular and renal endpoints, for which SGLT2i treatment is recommended in high risk groups in European/American guidelines.(1) Trial data provide an opportunity to explore the utility of non-routine features for treatment selection, which we did not explore as our focus was on developing a model for potential deployment in clinical practice in the near future. Evaluation of non-routine features in trial data, and genetic data, will be of particular interest for future study of underlying mechanisms of heterogeneous drug action.(45)

By providing validated patient level estimates of differences in glucose-lowering efficacy for these two major type 2 diabetes treatments, as well as assessing weight and discontinuation outcomes, our study has implications for clinical practice. The clinical features required to provide these estimates will be routinely measured and available to the majority of health professionals in developed countries. Work is ongoing to extend the prototype treatment selection decision aid to other type 2 diabetes treatment options; we have previously demonstrated differential treatment effects for sulfonylurea, thiazolidinedione, and GLP-1 receptor agonist therapy based on routine clinical features.(12, 46, 47)

## Conclusion

Simple clinical features can identify a large group of people with type 2 diabetes with a likely marked glycaemic benefit on SGLT2i therapy, and no increased risk of discontinuing treatment with SGLT2i compared with DPP4i. A smaller group have perhaps a clinically relevant glycaemic benefit on DPP4i, and meaningfully lower discontinuation risk on DPP4i compared with SGLT2i. These findings demonstrate the potential for a type 2 diabetes precision medicine approach based on routine features to inform clinical decisions concerning the choice of optimal glucose-lowering treatments.

## Supporting information

TRIPOD checklist

## Data Availability

No additional data are available from the authors although CPRD data are available by application to CPRD Independent Scientific Advisory Committee, and the clinical trial data are accessible via application from the Yale University Open Data Access Project and Vivli.

## Acknowledgements

This article is based in part on data from the Clinical Practice Research Datalink obtained under licence from the UK Medicines and Healthcare products Regulatory Agency. CPRD data is provided by patients and collected by the NHS as part of their care and support. This publication is based on research using data from data contributor Boehringer Ingelheim that has been made available through Vivli, Inc. Vivli has not contributed to or approved, and is not in any way responsible for, the contents of this publication. This study, carried out under YODA Project # 2017-1816, used data obtained from the Yale University Open Data Access Project, which has an agreement with JANSSEN RESEARCH & DEVELOPMENT, L.L.C.. The interpretation and reporting of research using this data are solely the responsibility of the authors and does not necessarily represent the official views of the Yale University Open Data Access Project or JANSSEN RESEARCH & DEVELOPMENT, L.L.C..

## Ethics/data approvals

Approval for the study was granted by the CPRD Independent Scientific Advisory Committee (ISAC 13_177R), the YODA Project (# 2017-1816) and Vivli (ID 00005959).

## Patient consent

Not applicable

## Contributor and guarantor information

The study concept and design were conceived and developed by JMD, AGJ, BMS, APM, and ATH. JMD and KGY undertook the analysis, with support from BMS. MDS developed the web calculator. All authors provided support for the analysis and interpretation of results, critically revised the article, and approved the final article. BMS and AGJ attest that all listed authors meet authorship criteria and that no others meeting the criteria have been omitted.

## Competing interests declaration

All authors have completed the ICMJE uniform disclosure form at www.icmje.org/coi_disclosure.pdf and declare:

APM declares previous research funding from Eli Lilly and Company, Pfizer, and AstraZeneca. BAM is an employee of the Wellcome Trust and holds an honorary post at University College London for the purposes of carrying out independent research; the views expressed in this manuscript do not necessarily reflect the views of the Wellcome Trust. SJV declares funding from IQVIA. WEH declares a grant from IQVIA. ERP declares personal fees from Sanofi and Lilly. RRH reports research support from AstraZeneca, Bayer and Merck Sharp & Dohme, and personal fees from Anji Pharmaceuticals, Bayer, Novartis and Novo Nordisk. NS declares personal fees from Abbott Diagnostics, Amgen, Astra Zeneca, Boehringer Ingelheim, Eli Lilly, Hanmi Pharmaceuticals, Novartis, Novo Nordisk, Pfizer and Sanofi and grants to his University from AstraZeneca, Boehringer Ingelheim, Novartis and Roche Diagnostics. Representatives from GSK, Takeda, Janssen, Quintiles, AstraZeneca and Sanofi attend meetings as part of the industry group involved with the MASTERMIND consortium. No industry representatives were involved in the writing of the manuscript or analysis of data. For all authors these are outside the submitted work; no other relationships or activities that could appear to have influenced the submitted work.

## Funding

This research was supported by a BHF-Turing Cardiovascular Data Science Award (SP/19/6/34809), and the Medical Research Council (UK) (MR/N00633X/1). JMD is supported by an Independent Fellowship funded by Research England’s Expanding Excellence in England (E3) fund. ATH and BMS are supported by the NIHR Exeter Clinical Research Facility. AGJ is supported by an NIHR Clinician Scientist fellowship (CS-2015-15-018). WH is supported by the NIHR Applied Research Collaboration South West Peninsula. SJV and BAM are supported by The Alan Turing Institute (EPSRC grant EP/N510129/). SJV is supported by the University of Warwick IAA funding. The views expressed are those of the authors and not necessarily those of the NHS, the NIHR or the Department of Health. The funders had no role in any part of the study or in any decision about publication.

## Transparency statement

The lead authors affirm that this manuscript is an honest, accurate, and transparent account of the study being reported; that no important aspects of the study have been omitted; and that any discrepancies from the study as planned have been explained.

## Copyright/license for publication

The Corresponding Author has the right to grant on behalf of all authors and does grant on behalf of all authors, a worldwide licence to the Publishers and its licensees in perpetuity, in all forms, formats and media (whether known now or created in the future), to i) publish, reproduce, distribute, display and store the Contribution, ii) translate the Contribution into other languages, create adaptations, reprints, include within collections and create summaries, extracts and/or, abstracts of the Contribution, iii) create any other derivative work(s) based on the Contribution, iv) to exploit all subsidiary rights in the Contribution, v) the inclusion of electronic links from the Contribution to third party material where-ever it may be located; and, vi) licence any third party to do any or all of the above.

**Tables: none**

## Notes

### Summary of Updates

Weblink for the research-only web tool implementing the algorithm developed for this study updated to: http://diabetes-calculator.uksouth.cloudapp.azure.com/calculator/

