## Supplementary Tables 1 for "Derivation and validation of a type 2 diabetes treatment selection algorithm for SGLT2-inhibitor and DPP4-inhibitor therapies based on glucose-lowering efficacy: cohort study using trial and routine clinical data"

### Supplementary material for: Derivation and validation of an algorithm to individualise type 2 diabetes treatment selection with SGLT2-inhibitor and DPP4-inhibitor therapy: prospective cohort study using trial and routine data

**Authors:** John M Dennis, Katherine G Young, Andrew P McGovern, Bilal A Mateen, Sebastian J Vollmer, Michael D Simpson, William E Henley, Rury R Holman, Naveed A Sattar, Ewan R Pearson, Andrew T Hattersley, Angus G Jones, Beverley M Shields

**sFlowchart: CPRD patient flow and inclusion criteria.**

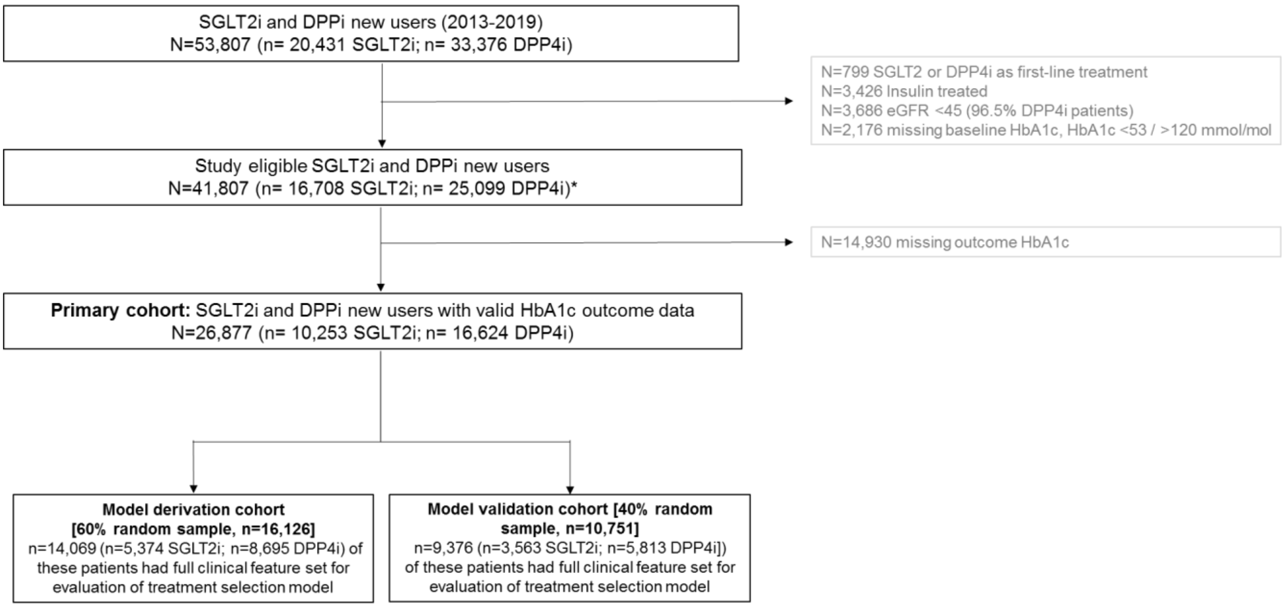


Baseline HbA1c defined as the closest HbA1c to drug initiation with –91/+7 days.

*For assessment of 6 month weight change, we followed the same procedures but included all patients with a baseline weight measure (within the 2 years prior to drug initiation) and outcome weight (closest weight to 6 month within 3-15 months, on unchanged glucose-lowering therapy), irrespective of whether they had an outcome HbA1c (n=20,065 [n= 7,842 SGLT2i, n=12,223 DPP4i). For assessment of treatment discontinuation, we included all patients with either 6 months of follow up time after drug initiation available, or 3 months of follow up time available after their last prescription (to confirm that the drug of interest was discontinued) (n=28,514 [n= 11,092 SGLT2i, n=17,422 DPP4i), irrespective of whether they had an outcome HbA1c

**sTable 1: Baseline clinical characteristics by initiated drug class.** Data are mean (SD) unless stated.

#### a) CPRD routine clinical data (primary cohort, n=26,877)

| | **DPP4-inhibitor (n=16,624)** | **SGLT2i (n=10,253)** | **SMD*** |
|---|---|---|---|
| Age (years) [mean SD] | 63.9 (10.8) | 60.0 (9.2) | 0.387 |
| Duration of diabetes (years) [mean SD] | 8.0 (5.3) | 8.5 (5.1) | 0.106 |
| **Sex** | | | |
| Female | 6265 (37.7) | 3751 (36.6) | 0.023 |
| Male | 10359 (62.3) | 6502 (63.4) | |
| **DPP4i type** | | | |
| Alogliptin | 1957 (11.8) | | |
| Linagliptin | 3331 (20.0) | | |
| Saxagliptin | 2207 (13.3) | | |
| Sitagliptin | 9025 (54.3) | | |
| Vildagliptin | 104 (0.6) | | |
| **SGLT2i type** | | | |
| Canaglifozin | | 1588 (15.5) | |
| Dapagliflozin | | 5861 (57.2) | |
| Empagliflozin | | 2804 (27.3) | |
| **Number of glucose-lowering drug classes ever prescribed** | | | |
| 1 | 7355 (44.2) | 2198 (21.4) | 0.547 |
| 2 | 7024 (42.3) | 3187 (31.1) | |
| 3 | 1901 (11.4) | 3027 (29.5) | |
| 4+ | 344 (2.1) | 1841 (18.0) | |
| **Number of current other glucose-lowering drugs** | | | |
| 0 | 947 (5.7) | 286 (2.8) | 0.844 |
| 1 | 9702 (58.4) | 4117 (40.2) | |
| 2 | 5770 (34.7) | 4701 (45.8) | |
| 3+ | 205 (1.2) | 1149 (11.2) | |
| **Baseline biomarkers (all mean SD)** | | | |
| HbA1c (mmol/mol) | 73.0 (13.4) | 76.9 (14.2) | 0.285 |
| BMI (kg/m$^2$) | 32.3 (6.4) | 34.4 (6.6) | 0.312 |
| eGFR (mL/min/1.3 m$^2$) | 83.1 (17.3) | 88.8 (14.7) | 0.356 |
| HDL-c (mmol/L) | 1.2 (0.3) | 1.1 (0.3) | 0.151 |
| Triglycerides (mmol/L) | 2.2 (1.6) | 2.4 (1.6) | 0.078 |
| ALT (IU/L) | 33.7 (52.4) | 36.6 (40.2) | 0.061 |
| Albumin (g/L) | 42.2 (4.0) | 42.2 (4.0) | 0.014 |
| Bilurubin (µmol/L) | 10.0 (5.1) | 9.8 (5.0) | 0.040 |
| **HbA1c outcome** | | | |
| Outcome HbA1c (mmol/mol) [mean SD] | 65.1 (16.1) | 65.1 (14.4) | 0.001 |
| Month of outcome HbA1c measure | 9.2 (3.4) | 9.0 (3.5) | 0.050 |

*Standardized mean difference: ≥0.1 is a metric for a meaningful imbalance.

#### b) Clinical trials

| | CANTATA-D | | CANTATA-D2 | | EMPA-REG MONO | |
|---|---|---|---|---|---|---|
| | **SGLT2-inhibitor (n=705) [Canagliflozin]** | **DPP4-inhibitor (n=355) [Sitagliptin]** | **SGLT2-inhibitor (n=359) [Canagliflozin]** | **DPP4-inhibitor (n=356) [Sitagliptin]** | **SGLT2-inhibitor (n=505) [Empagliflozin]** | **DPP4-inhibitor (n=219) [Sitagliptin]** |
| **Age (years)** | 55.2 (9.3) | 55.4 (9.5) | 56.5 (9.6) | 56.6 (9.3) | 54.3 (11.6) | 55.1 (9.9) |
| **Sex** | | | | | | |
| Female | 382 (54.2) | 187 (52.7) | 161 (44.8) | 151 (43.4) | 173 (34.3) | 82 (37.4) |
| Male | 323 (45.8) | 168 (47.3) | 198 (55.2) | 205 (57.6) | 332 (65.7) | 137 (62.6) |
| **HbA1c (mmol/mol)** | 63.3 (9.9) | 63.1 (9.5) | 65.1 (9.9) | 65.4 (10.0) | 68.2 (16.9) | 62.2 (8.5) |
| **BMI (kg/m$^2$)** | 31.9 (6.4) | 32.0 (6.1) | 31.5 (6.9) | 31.7 (6.9) | 28.3 (5.5) | 28.2 (5.2) |
| **eGFR (mL/min/1.3 m$^2$)** | 89.3 (18.2) | 88.1 (18.8) | 87.1 (18.1) | 87.7 (20.3) | 88.6 (19.2) | 87.6 (17.4) |
| **HDL-c (mmol/L)** | 1.2 (0.3) | 1.2 (0.3) | 1.2 (0.3) | 1.2 (0.3) | 1.2 (0.3) | 1.3 (0.4) |
| **Triglycerides (mmol/L)** | 2.1 (1.5) | 2.0 (1.1) | 2.0 (1.4) | 1.9 (1.3) | 2.2 (2.4) | 2.2 (2.0) |
| **ALT (IU/L)** | 29.1 (19.6) | 28.5 (13.9) | 27.8 (13.5) | 27.8 (15.5) | 31.4 (17.4) | 31.9 (15.2) |
| **Albumin (g/L)** | 41.0 (3.1) | 41.1 (3.3) | 40.9 (3.4) | 40.9 (3.2) | 45.2 (3.0) | 45.5 (2.6) |
| **Bilirubin (μmol/L)** | 8.7 (4.2) | 8.8 (4.4) | 8.2 (4.2) | 8.3 (4.1) | 10.0 (4.8) | 9.8 (4.8) |
| **Background therapy** | | | | | | |
| Metformin | 705 (100.0) | 355 (100.0) | 359 (100.0) | 356 (100.0) | 0 (0.0) | 0 (0.0) |
| Sulfonylurea | 0 (0.0) | 0 (0.0) | 359 (100.0) | 356 (100.0) | 0 (0.0) | 0 (0.0) |
| Thiazolidinedione | 0 (0.0) | 0 (0.0) | 0 (0.0) | 0 (0.0) | 0 (0.0) | 0 (0.0) |
| DPP4-inhibitor | 0 (0.0) | 0 (0.0) | 0 (0.0) | 0 (0.0) | 0 (0.0) | 0 (0.0) |
| Other* | 0 (0.0) | 0 (0.0) | 0 (0.0) | 0 (0.0) | 0 (0.0) | 0 (0.0) |

*other = α-glucosidase inhibitors, glucagon-like peptide-1 agonists, glinides

Values shown are mean (standard deviation) or n (%) for participants included in the mixed effects models and subsequent meta-analysis. This includes participants who were not insulin-treated and with at least one on-treatment HbA1c between randomisation and 6 months. EMPA-REG MONO/PIO/METSU: HbA1c measurements excluded after changes in study medication (including dose changes); EMPA-REG OUTCOME: HbA1c measurements excluded after changes in study medication (including dose changes) or changes in background medication (not including dose changes). Study medication (trial arm) is as randomised in all CANTATA trials, and as per actual study medication given in all other trials.

BMI, body mass index; DPP4-inhibitor, dipeptidyl-peptidase 4 inhibitor; eGFR, estimated glomerular filtration rate; HbA1c, glycated haemoglobin, type A1c; HDL-c, high-density lipoprotein cholesterol; SGLT2-inhibitor, sodium-glucose cotransporter-2 inhibitor.

| | CANTATA-SU (n=939) [Canagliflozin] | CANTATA-M (n=459) [Canagliflozin] | NCT01106651 (n=303) [Canagliflozin] | CANTATA-MSU (n=303) [Canagliflozin] | EMPA-REG PIO (n=321) [Empagliflozin] | EMPA-REG METSU (n=1,015) [Empagliflozin] |
|---|---|---|---|---|---|---|
| **Age (years)** | 55.9 (9.3) | 53.8 (10.8) | 64.1 (6.3) | 56.7 (9.8) | 54.5 (9.5) | 55.5 (10.0) |
| **Sex** | | | | | | |
| Female | 460 (49.0) | 257 (56.0) | 138 (45.5) | 142 (46.9) | 161 (50.2) | 456 (44.9) |
| Male | 479 (51.0) | 202 (44.0) | 165 (54.5) | 161 (53.1) | 160 (49.8) | 559 (55.1) |
| **HbA1c (mmol/mol)** | 61.5 (8.5) | 69.5 (15.2) | 60.3 (8.6) | 65.2 (10.1) | 64.7 (9.4) | 69.6 (16.3) |
| **BMI (kg/m$^2$)** | 31.1 (5.3) | 31.3 (6.3) | 31.1 (4.5) | 33.3 (6.3) | 29.2 (5.6) | 29.0 (5.5) |
| **eGFR (mL/min/1.3 m$^2$)** | 90.6 (19.3) | 88.5 (19.8) | 77.8 (16.7) | 89.9 (19.8) | 85.8 (22.6) | 89.2 (21.1) |
| **HDL-c (mmol/L)** | 1.2 (0.3) | 1.2 (0.3) | 1.2 (0.3) | 1.1 (0.3) | 1.3 (0.3) | 1.3 (0.3) |
| **Triglycerides (mmol/L)** | 2.1 (1.8) | 2.1 (1.3) | 1.8 (1.1) | 2.2 (1.5) | 1.8 (1.7) | 1.9 (1.3) |
| **ALT (IU/L)** | 28.9 (15.9) | 28.6 (15.9) | 26.2 (13.0) | 28.9 (13.5) | 23.6 (11.1) | 28.2 (15.9) |
| **Albumin (g/L)** | 42.2 (3.2) | 41.1 (3.3) | 40.9 (2.8) | 40.9 (3.0) | 44.6 (2.8) | 45.2 (2.9) |
| **Bilirubin (μmol/L)** | 8.8 (4.0) | 9.4 (4.5) | 8.7 (3.9) | 8.7 (5.0) | 8.7 (4.3) | 9.2 (4.5) |
| **Background therapy** | | | | | | |
| Metformin | 939 (100.0) | 0 (0.0) | 278 (91.7) | 303 (100.0) | 245 (76.3) | 1,015 (100.0) |
| Sulfonylurea | 0 (0.0) | 0 (0.0) | 183 (60.5) | 303 (100.0) | 0 (0.0) | 521 (51.3) |
| Thiazolidinedione | 0 (0.0) | 0 (0.0) | 42 (13.7) | 0 (0.0) | 321 (100.0) | 0 (0.0) |
| DPP4-inhibitor | 0 (0.0) | 0 (0.0) | 32 (10.5) | 0 (0.0) | 0 (0.0) | 0 (0.0) |
| Other* | 0 (0.0) | 0 (0.0) | 17 (5.7) | 0 (0.0) | 0 (0.0) | 0 (0.0) |

*other = α-glucosidase inhibitors, glucagon-like peptide-1 agonists, glinides

Values shown are mean (standard deviation) or n (%) for participants included in the mixed effects models and subsequent meta-analysis. This includes participants who were not insulin-treated and with at least one on-treatment HbA1c between randomisation and 6 months. EMPA-REG MONO/PIO/METSU: HbA1c measurements excluded after changes in study medication (including dose changes); EMPA-REG OUTCOME: HbA1c measurements excluded after changes in study medication (including dose changes) or changes in background medication (not including dose changes). Study medication (trial arm) is as randomised in all CANTATA trials, and as per actual study medication given in all other trials.

BMI, body mass index; DPP4-inhibitor, dipeptidyl-peptidase 4 inhibitor; eGFR, estimated glomerular filtration rate; HbA1c, glycated haemoglobin, type A1c; HDL-c, high-density lipoprotein cholesterol; SGLT2-inhibitor, sodium-glucose cotransporter-2 inhibitor.

| | EMPA-REG OUTCOME (n=2,210) [Empagliflozin] | NCT00602472 (n=774) [Linagliptin] | NCT00621140 (n=326) [Linagliptin] | NCT00601250 (n=506) [Linagliptin] | NCT00622284 (n=759) [Linagliptin] |
|---|---|---|---|---|---|
| **Age (years)** | 62.7 (8.8) | 58.3 (9.9) | 56.3 (10.1) | 56.6 (10.0) | 59.8 (9.4) |
| **Sex** | | | | | |
| Female | 596 (27.0) | 411 (53.1) | 167 (51.2) | 238 (47.0) | 310 (40.8) |
| Male | 1,614 (73.0) | 363 (46.9) | 159 (48.8) | 268 (53.0) | 449 (59.2) |
| **HbA1c (mmol/mol)** | 63.4 (9.3) | 65.5 (8.8) | 63.7 (9.4) | 64.8 (9.3) | 60.5 (9.6) |
| **BMI (kg/m$^2$)** | 29.7 (5.1) | 28.4 (4.8) | 29.0 (4.8) | 29.9 (4.8) | 30.2 (4.8) |
| **eGFR (mL/min/1.3 m$^2$)** | 76.8 (20.5) | 95.8 (22.9) | 90.1 (22.4) | 101.2 (29.9) | 93.0 (22.5) |
| **HDL-c (mmol/L)** | 1.2 (0.3) | 1.2 (0.3) | 1.2 (0.4) | 1.2 (0.3) | 1.2 (0.3) |
| **Triglycerides (mmol/L)** | 1.9 (1.4) | 2.1 (2.1) | 1.9 (1.1) | 2.0 (1.4) | 1.9 (1.4) |
| **ALT (IU/L)** | 26.1 (14.1) | 27.2 (15.8) | 25.7 (15.3) | 27.2 (15.5) | 27.8 (15.9) |
| **Albumin (g/L)** | 44.8 (2.9) | 45.3 (2.7) | 45.1 (3.0) | 44.8 (2.5) | 44.8 (2.4) |
| **Bilirubin (µmol/L)** | 8.6 (4.6) | 9.3 (4.3) | 10.7 (5.8) | 9.0 (4.7) | 9.0 (4.7) |
| **Background therapy** | | | | | |
| Metformin | 1,819 (82.3) | 774 (100.0) | 0 (0.0) | 506 (100.0) | 759 (100.0) |
| Sulfonylurea | 1,419 (64.2) | 774 (100.0) | 0 (0.0) | 0 (0.0) | 0 (0.0) |
| Thiazolidinedione | 125 (5.7) | 0 (0.0) | 0 (0.0) | 0 (0.0) | 0 (0.0) |
| DPP4-inhibitor | 324 (14.7) | 0 (0.0) | 0 (0.0) | 0 (0.0) | 0 (0.0) |
| Other* | 225 (10.2) | 0 (0.0) | 0 (0.0) | 0 (0.0) | 0 (0.0) |

*other = α-glucosidase inhibitors, glucagon-like peptide-1 agonists, glinides

Values shown are mean (standard deviation) or n (%) for participants included in the mixed effects models and subsequent meta-analysis. This includes participants who were not insulin-treated and with at least one on-treatment HbA1c between randomisation and 6 months. EMPA-REG MONO/PIO/METSU: HbA1c measurements excluded after changes in study medication (including dose changes); EMPA-REG OUTCOME: HbA1c measurements excluded after changes in study medication (including dose changes) or changes in background medication (not including dose changes). Study medication (trial arm) is as randomised in all CANTATA trials, and as per actual study medication given in all other trials.

BMI, body mass index; DPP4-inhibitor, dipeptidyl-peptidase 4 inhibitor; eGFR, estimated glomerular filtration rate; HbA1c, glycated haemoglobin, type A1c; HDL-c, high-density lipoprotein cholesterol; SGLT2-inhibitor, sodium-glucose cotransporter-2 inhibitor.

**sTable 2: Associations between clinical features and baseline HbA1c adjusted 6 month HbA1c response for SGLT2-inhibitor and DPP4-inhibitor treatment**

a) **CPRD routine clinical data (primary cohort, n=26,877). Data underlying Figure 2a.** Estimates are derived from separate models for each clinical feature. N's represent the number of patients included in the model for each clinical feature. Each model was adjusted for baseline HbA1c-by-drug interaction, number of glucose-lowering drug classes ever prescribed, number of current glucose-lowering treatments, and month of HbA1c outcome measurement.

| | SGLT2-inhibitor | DPP4-inhibitor |
|---|---|---|
| | Beta coefficient (95%CI) | Beta coefficient (95%CI) |
| Male Sex (n=26,877) | -1.39 (-1.92;-0.86) | -0.40 (-0.82;0.02) |
| Current age per year (n=26,877) | 0.27 (-0.02;0.56) | -1.86 (-2.05;-1.66) |
| Diabetes Duration per year (n=26,877) | 0.41 (0.12;0.70) | -1.12 (-1.33;-0.91) |
| eGFR per 1 mL/min/1.3 $m^2$ (n=26,877) | -0.81 (-1.10;-0.52) | 1.45 (1.25;1.64) |
| BMI per 1 kg/$m^2$ (n=26,877) | -0.21 (-0.47;0.05) | 0.96 (0.75;1.17) |
| HDL-c per 1 mmol/L (n=26,877) | 0.62 (0.36;0.89) | -0.39 (-0.60;-0.19) |
| Triglycerides per 1 mmol/L (n=26,877) | 0.05 (-0.24;0.33) | 0.99 (0.78;1.21) |
| ALT per 1 IU/L (n=26,877) | -1.42 (-1.70;-1.14) | 0.57 (0.35;0.78) |
| Albumin per 1 g/L (n=26,877) | -0.59 (-0.85;-0.32) | 0.16 (-0.05;0.37) |
| Bilurubin per 1 µmol/L (n=26,877) | -1.01 (-1.27;-0.75) | -0.40 (-0.61;-0.20) |

**Clinical trial data (meta-analysis of 14 clinical trials). Data underlying Figure 2b.** For each clinical feature, estimates are derived from random effects meta-analysis of beta coefficients estimated separately in each trial. $I^2$ represents the fraction of variance that is due to heterogeneity across the trials. N's represent the number of patients in the included in the model for each clinical feature.

#### a) Meta-analysis

| | SGLT2-inhibitor | | | DPP4-inhibitor | | |
|---|---|---|---|---|---|---|
| | **Beta coefficient (95%CI)** | **$I^2$** | **N** | **Beta coefficient (95%CI)** | **$I^2$** | **N** |
| **Male Sex** | -1.05 (-1.63;-0.47) | 70.3% | 7,119 | -0.20 (-0.60;0.20) | 0.0% | 3,295 |
| **Current age per SD change** | 0.76 (0.60;0.92) | 0.0% | 7,117 | 0.12 (-0.10;0.33) | 0.8% | 3,295 |
| **Diabetes Duration per SD change** | NA | NA | NA | NA | NA | NA |
| **eGFR per SD change** | -0.82 (-0.96;-0.68) | 1.04% | 7,104 | 0.01 (-0.16;0.18) | 0.0% | 3,226 |
| **BMI per SD change** | 0.09 (-0.14;0.33) | 40.7% | 7,116 | 0.70 (0.41;0.99) | 21.3% | 3,295 |
| **HDL-c per SD change** | 0.55 (0.38;0.73) | 23.5% | 7,054 | -0.08 (-0.26;0.11) | 0.0% | 3,166 |
| **Triglycerides per SD change** | 0.15 (0.00;0.29) | 0.0% | 7,067 | 0.56 (0.36;0.76) | 0.0% | 3,166 |
| **ALT per SD change** | -0.79 (-1.02;-0.55) | 43.1% | 7,027 | 0.43 (0.13;0.73) | 40.4% | 3,130 |
| **Albumin per SD change** | -0.84 (-1.12;-0.56) | 47.0% | 7,079 | -0.57 (-0.86,-0.28) | 0.0% | 3,184 |
| **Bilurubin per SD change** | -0.52 (-0.74;-0.29) | 36.2% | 6,981 | -0.26 (-0.65;0.13) | 65.6% | 3,166 |

#### b) Individual trials

| | CANTATA-D | | | | | | CANTATA-D2 | | | | | | EMPA-REG MONO | | | | | |
|---|---|---|---|---|---|---|---|---|---|---|---|---|---|---|---|---|---|---|
| | SGLT2-inhibitor (n=705) [Canagliflozin] | | | DPP4-inhibitor (n=355) [Sitagliptin] | | | SGLT2-inhibitor (n=359) [Canagliflozin] | | | DPP4-inhibitor (n=356) [Sitagliptin] | | | SGLT2-inhibitor (n=505) [Empagliflozin] | | | DPP4-inhibitor (n=219) [Sitagliptin] | | |
| | **Beta** | **SE** | **n** | **Beta** | **SE** | **n** | **Beta** | **SE** | **n** | **Beta** | **SE** | **n** | **Beta** | **SE** | **n** | **Beta** | **SE** | **n** |
| **Male sex** | -0.120 | 0.460 | 705 | 0.634 | 0.641 | 355 | -0.978 | 0.648 | 359 | -0.160 | 0.652 | 356 | -1.639 | 0.714 | 505 | -0.785 | 0.822 | 219 |
| **Current age** | 0.728 | 0.254 | 705 | -0.149 | 0.347 | 355 | 0.720 | 0.348 | 359 | -0.008 | 0.369 | 356 | 0.802 | 0.308 | 504 | 0.030 | 0.418 | 219 |
| **eGFR** | -0.826 | 0.236 | 702 | -0.053 | 0.321 | 353 | -0.539 | 0.344 | 356 | -0.062 | 0.304 | 354 | -0.424 | 0.334 | 504 | 0.482 | 0.433 | 219 |
| **BMI** | 0.260 | 0.232 | 703 | 0.464 | 0.345 | 355 | 0.618 | 0.300 | 359 | 0.566 | 0.306 | 356 | -0.316 | 0.403 | 505 | 1.288 | 0.496 | 219 |
| **HDL-c** | 0.316 | 0.229 | 700 | 0.202 | 0.358 | 353 | 0.756 | 0.315 | 358 | 0.210 | 0.315 | 354 | 1.271 | 0.287 | 498 | 0.026 | 0.347 | 218 |
| **Triglycerides** | 0.086 | 0.217 | 704 | 0.724 | 0.341 | 354 | -0.296 | 0.298 | 358 | 0.556 | 0.320 | 355 | -0.270 | 0.291 | 498 | 0.376 | 0.351 | 218 |
| **ALT** | -0.577 | 0.245 | 699 | -0.245 | 0.377 | 351 | -0.332 | 0.382 | 354 | 1.029 | 0.356 | 354 | -0.694 | 0.364 | 500 | 0.473 | 0.454 | 218 |
| **Albumin** | -0.741 | 0.291 | 699 | -0.286 | 0.393 | 351 | -0.675 | 0.378 | 355 | -0.280 | 0.396 | 354 | -0.342 | 0.457 | 502 | -1.717 | 0.600 | 218 |
| **Bilirubin** | 0.016 | 0.282 | 684 | -0.469 | 0.382 | 346 | -0.265 | 0.401 | 343 | 0.795 | 0.413 | 344 | -0.870 | 0.356 | 502 | -1.335 | 0.410 | 218 |

| | CANTATA-SU (n=939) [Canagliflozin] | | | CANTATA-M (n=459) [Canagliflozin] | | | NCT01106651 (n=303) [Canagliflozin] | | | CANTATA-MSU (n=303) [Canagliflozin] | | | EMPA-REG PIO (n=321) [Empagliflozin] | | | EMPA-REG METSU (n=1,015) [Empagliflozin] | | |
|---|---|---|---|---|---|---|---|---|---|---|---|---|---|---|---|---|---|---|
| | **Beta** | **SE** | **n** | **Beta** | **SE** | **n** | **Beta** | **SE** | **n** | **Beta** | **SE** | **n** | **Beta** | **SE** | **n** | **Beta** | **SE** | **n** |
| **Male sex** | -1.267 | 0.371 | 939 | 0.034 | 0.624 | 459 | -0.619 | 0.567 | 303 | -0.953 | 0.596 | 303 | 0.123 | 0.757 | 321 | -1.949 | 0.415 | 1,015 |
| **Current age** | 0.599 | 0.206 | 939 | 0.907 | 0.298 | 459 | 0.806 | 0.463 | 303 | 0.644 | 0.314 | 303 | 1.000 | 0.411 | 321 | 0.696 | 0.218 | 1,015 |
| **eGFR** | -0.671 | 0.183 | 937 | -1.042 | 0.302 | 457 | -0.390 | 0.321 | 301 | -0.610 | 0.281 | 301 | -0.962 | 0.313 | 321 | -0.809 | 0.187 | 1,015 |
| **BMI** | -0.380 | 0.228 | 939 | 0.018 | 0.325 | 459 | 0.214 | 0.414 | 302 | -0.050 | 0.305 | 303 | 0.497 | 0.447 | 321 | 0.519 | 0.248 | 1,015 |
| **HDL-c** | 0.702 | 0.178 | 935 | 0.252 | 0.323 | 454 | 0.437 | 0.298 | 302 | 0.528 | 0.320 | 302 | 0.090 | 0.361 | 320 | 0.621 | 0.198 | 1,001 |
| **Triglycerides** | 0.130 | 0.176 | 938 | 0.324 | 0.303 | 459 | 0.230 | 0.303 | 303 | 0.138 | 0.270 | 302 | 0.595 | 0.381 | 320 | -0.252 | 0.203 | 1,001 |
| **ALT** | -0.844 | 0.203 | 929 | -0.702 | 0.355 | 454 | -0.429 | 0.344 | 301 | -0.396 | 0.345 | 300 | -1.043 | 0.450 | 319 | -0.900 | 0.227 | 997 |
| **Albumin** | -0.626 | 0.234 | 932 | -0.296 | 0.376 | 455 | -0.674 | 0.401 | 301 | -0.731 | 0.393 | 301 | -0.759 | 0.548 | 320 | -1.634 | 0.285 | 1,004 |
| **Bilirubin** | -0.166 | 0.243 | 918 | -0.972 | 0.348 | 446 | -0.370 | 0.379 | 290 | -0.310 | 0.311 | 289 | -0.534 | 0.448 | 320 | -1.023 | 0.239 | 1,003 |

| | EMPA-REG OUTCOME (n=2,210) [Empagliflozin] | | | NCT00602472 (n=770) [Linagliptin] | | | NCT00621140 (n=326) [Linagliptin] | | | NCT00601250 (n=506) [Linagliptin] | | | NCT00622284 (n=759) [Linagliptin] | | |
|---|---|---|---|---|---|---|---|---|---|---|---|---|---|---|---|
| | **Beta** | **SE** | **n** | **Beta** | **SE** | **n** | **Beta** | **SE** | **n** | **Beta** | **SE** | **n** | **Beta** | **SE** | **n** |
| **Male sex** | -2.390 | 0.329 | 2,210 | 0.279 | 0.438 | 774 | -0.860 | 0.768 | 326 | -0.490 | 0.563 | 506 | -0.428 | 0.363 | 759 |
| **Current age** | 0.861 | 0.173 | 2,209 | -0.146 | 0.228 | 774 | -0.082 | 0.397 | 326 | 0.121 | 0.293 | 506 | 0.495 | 0.196 | 759 |
| **eGFR** | -1.061 | 0.134 | 2,210 | 0.117 | 0.182 | 744 | 0.320 | 0.333 | 311 | 0.048 | 0.183 | 486 | -0.174 | 0.150 | 759 |
| **BMI** | -0.187 | 0.187 | 2,210 | 1.099 | 0.292 | 774 | 1.331 | 0.519 | 326 | 0.254 | 0.379 | 506 | 0.476 | 0.243 | 759 |
| **HDL-c** | 0.444 | 0.148 | 2,184 | -0.362 | 0.212 | 751 | -0.099 | 0.303 | 298 | -0.044 | 0.269 | 473 | -0.067 | 0.171 | 719 |
| **Triglycerides** | -0.014 | 0.156 | 2,184 | 0.713 | 0.200 | 760 | 0.806 | 0.429 | 296 | 0.561 | 0.293 | 469 | 0.323 | 0.186 | 714 |
| **ALT** | -1.346 | 0.166 | 2,174 | 0.519 | 0.230 | 739 | 1.007 | 0.444 | 299 | 0.429 | 0.319 | 460 | 0.135 | 0.195 | 709 |
| **Albumin** | -1.286 | 0.202 | 2,210 | -0.659 | 0.307 | 762 | -0.331 | 0.529 | 308 | -0.861 | 0.436 | 470 | -0.455 | 0.300 | 721 |
| **Bilirubin** | -0.605 | 0.163 | 2,186 | -0.292 | 0.248 | 759 | -0.534 | 0.342 | 308 | -0.224 | 0.300 | 470 | 0.105 | 0.198 | 721 |

**Mixed effects model results used for the trial meta-analysis**

Beta, SE and n are the beta coefficients, standard error of the means and number of participants from mixed effects models of the association between individual clinical features and HbA1c outcome within each trial arm*. These models included patient-level random effects and adjusted for baseline HbA1c. Clinical features were standardised to CPRD means and standard deviations, so that beta coefficients represent the change in outcome HbA1c in mmol/mol per standard deviation change in the clinical feature (ALT and triglycerides values were logged prior to inclusion in the model). Diabetes duration was not available (except in categorised form) in any of the trial data.

Using notation from the nlme R package, the mixed models were of the form:
[outcome HbA1c] ~ factor([time point]) + [baseline HbA1c] + [clinical feature of interest] + factor([SGLT2-inhibitor dose]), random = ~ [time point] | [patient ID]

Outcome HbA1c: on-therapy HbA1c values up to 6 months post-randomisation. EMPA-REG MONO/PIO/METSU: HbA1c measurements excluded after changes in study medication (including dose changes); EMPA-REG OUTCOME: HbA1c measurements excluded after changes in study medication (including dose changes) or changes in background medication (not including dose changes).

Time point: time point post-randomisation at which the outcome HbA1c was measured (1 for the first time point post-randomisation in a trial, 2 for the second etc.).

Total number of observations per model was > n as individuals have multiple on-therapy HbA1c values up to 6 months post-randomisation). Participants who were insulin-treated were not included in the analysis.

*Study medication (trial arm) is as randomised in all CANTATA trials, and as per actual study medication given in all other trials.

BMI, body mass index; DPP4-inhibitor, dipeptidyl-peptidase 4 inhibitor; eGFR, estimated glomerular filtration rate; HbA1c, glycated haemoglobin, type A1c; HDL-c, high-density lipoprotein cholesterol; SGLT2-inhibitor, sodium-glucose cotransporter-2 inhibitor.

**sFigure 1: Relative feature importance for treatment selection between SGLT2-inhibitor and DPP4-inhibitor treatment, for all clinical features in multivariable analysis in routine clinical data.** Feature importance reflects the proportion of chi-squared explained by drug-by-covariate interaction terms for each clinical feature, as these represent differential treatment effects for the two therapies. Model estimated from complete case analysis of 12,034 CPRD patients with full clinical feature set available in the primary care record. Bars represent bootstrapped 95% confidence intervals.

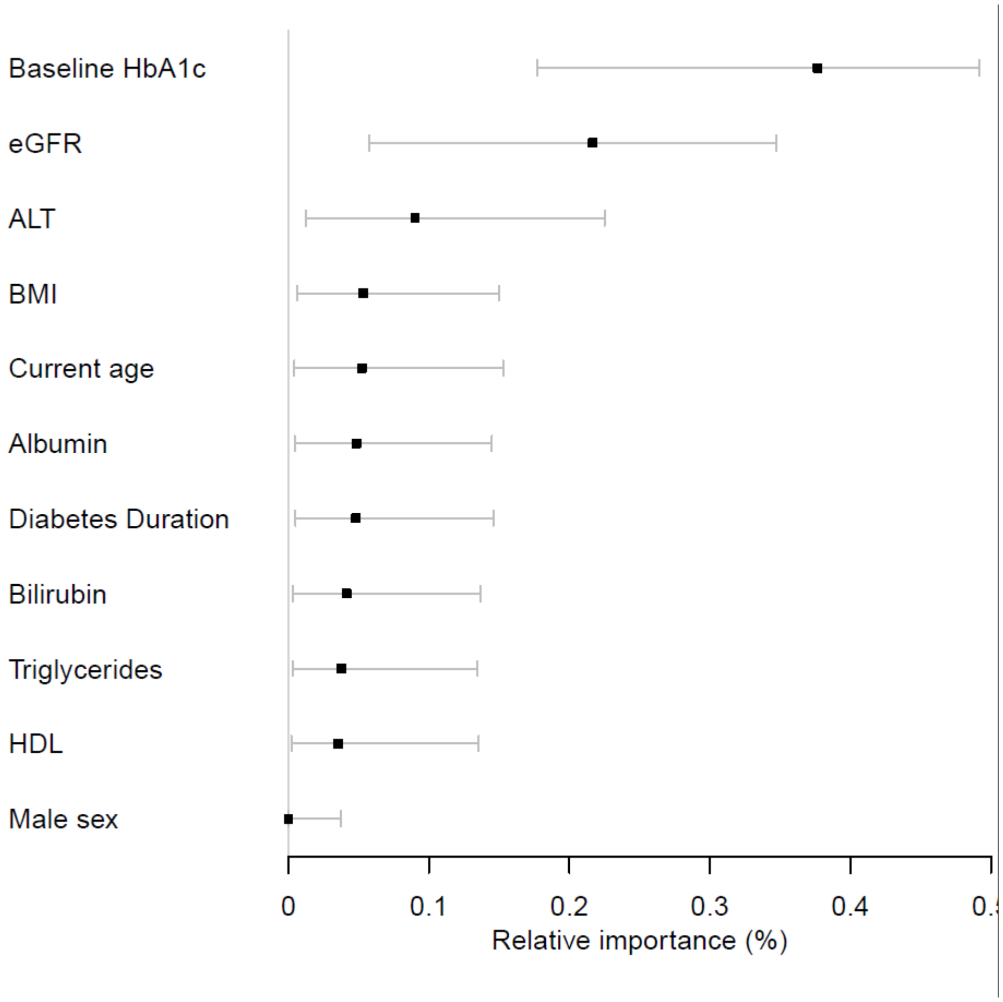

**sTable 3: Full treatment selection model equation.** Drugclass = “SGLT2” or “DPP4” (factor variable), prehba1cmmol = baseline HbA1c, drugline = number of current and previous therapies (including therapy initiated), ncurrtx = number of current glucose-lowering therapies, hba1cmonth = month of HbA1c outcome measure, egfr_ckdepi = eGFR (CKD-EPI equation), prealtlog = Alanine aminotransferase (logged), prebmi = BMI, agetx = current age. To predict achieved 6 month HbA1c for a patient initiating second-line treatment (e.g. after metformin), set drugline == 2, ncurrtx == 1, hba1cmonth == 6, and input patient-specific values of other clinical features.

| **Full model equation** |
|---|
| 31.321954+0.64204609* prehba1cmmol-8.5439347e-05*pmax(prehba1cmmol-59,0)^3+0.00012815908*pmax(prehba1cmmol-71.000008,0)^3-4.2719731e-05*pmax(prehba1cmmol-94.999992,0)^3+33.896175*(drugclass=="SGLT2")+4.3479453*(drugline=="3")+7.7707306*(drugline=="4")+9.8519056*(drugline=="5")-6.0952285*(ncurrtx=="1")-8.4474283*(ncurrtx=="2")-10.076309*(ncurrtx=="3")+0.01458388*egfr_ckdepi+6.5992202e-07*pmax(egfr_ckdepi-61.712658,0)^3-1.6584462e-06*pmax(egfr_ckdepi-87.730606,0)^3+9.9852416e-07*pmax(egfr_ckdepi-104.9258,0)^3-0.59830102* prealtlog+0.38060553*pmax(prealtlog-2.7080502,0)^3-0.75191225*pmax(prealtlog-3.3672958,0)^3+0.37130672*pmax(prealtlog-4.0430513,0)^3+0.090509535* prebmi-0.00016688487*pmax(prebmi-25.700001,0)^3+0.00027814137*pmax(prebmi-32.099998,0)^3-0.0001112565*pmax(prebmi-41.700001,0)^3-0.093145764* agetx-9.4518092e-05*pmax(agetx-49,0)^3+0.00018228489*pmax(agetx-62,0)^3-8.77668e-05*pmax(agetx-76,0)^3-0.62516011*hba1cmonth+0.0042770662*pmax(hba1cmonth-4,0)^3-0.012831199*pmax(hba1cmonth-10,0)^3+0.0085541324*pmax(hba1cmonth-13,0)^3+(drugclass=="SGLT2")*(-0.26397744*prehba1cmmol+0.0002001556*pmax(prehba1cmmol-59,0)^3-0.00030023353*pmax(prehba1cmmol-71.000008,0)^3+0.00010007793*pmax(prehba1cmmol-94.999992,0)^3)+(drugclass=="SGLT2")*(-0.10243175*egfr_ckdepi+1.8080869e-05*pmax(egfr_ckdepi-61.712658,0)^3-4.5438926e-05*pmax(egfr_ckdepi-87.730606,0)^3+2.7358057e-05*pmax(egfr_ckdepi-104.9258,0)^3)+(drugclass=="SGLT2")*(-2.4219094*prealtlog+0.24680001*pmax(prealtlog-2.7080502,0)^3-0.48757028*pmax(prealtlog-3.3672958,0)^3+0.24077028*pmax(prealtlog-4.0430513,0)^3)+(drugclass=="SGLT2")*(-0.25565408*prebmi+0.00099460814*pmax(prebmi-25.700001,0)^3-0.0016576797*pmax(prebmi-32.099998,0)^3+0.00066307158*pmax(prebmi-41.700001,0)^3)+(drugclass=="SGLT2")*(0.061244729*agetx+0.00010509417*pmax(agetx-49,0)^3-0.00020268161*pmax(agetx-62,0)^3+9.758744e-05*pmax(agetx-76,0)^3) |

**sFigure 2: Visualisation of non-linear associations for continuous clinical variables in the full multivariable treatment selection model (as reported in sTable 3).** Predictions are scaled to: baseline HbA1c 58, mean value of other continuous covariates and baseline defaults of other features: number of current and previous therapies (including therapy initiated) = 2, number of current glucose-lowering therapies = 1, month of HbA1c outcome measure = 6, eGFR = 87.9, Alanine aminotransferase (logged) = 3.33, BMI = 32.1, current age = 62.

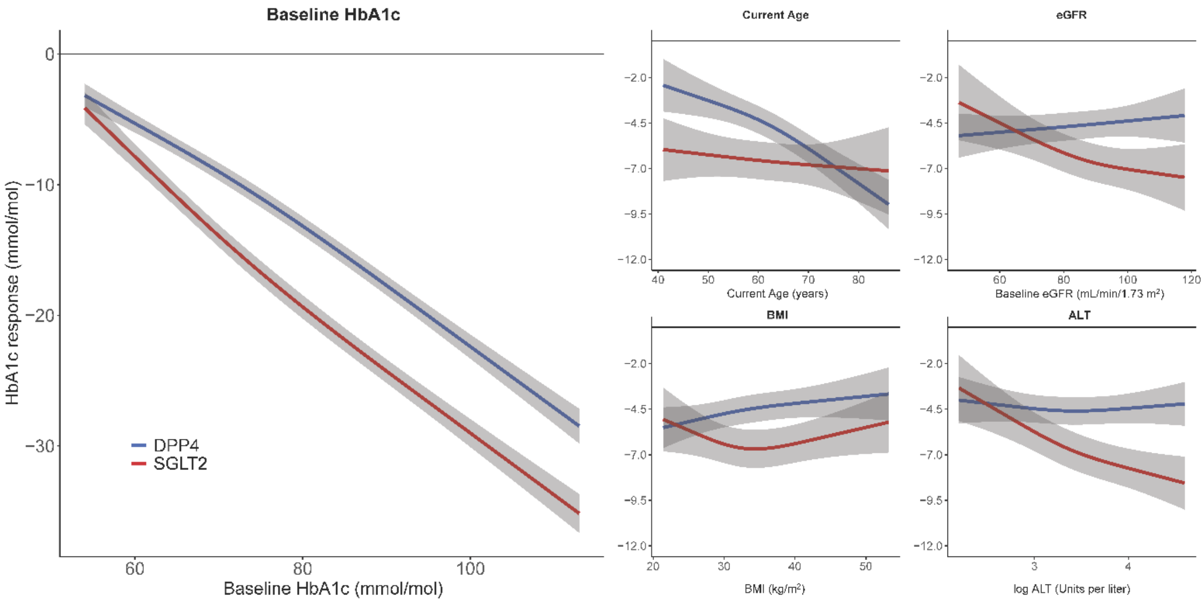

**sTable 4: Model performance statistics for predicting HbA1c outcome**

**a) Internal validation (CPRD derivation cohort, n=14,069).** Model outcome is 6 month HbA1c.
*Optimism corrected performance estimated from 1000 bootstraps with replacement

| | Apparent performance (95%CI) | Average optimism* | Final model optimism corrected performance (95%CI)* |
|---|---|---|---|
| **$R^2$** | 0.294 (0.278, 0.310) | 0.0026 | 0.29 (0.27, 0.31) |
| **Root mean square error (mmol/mol)** | 12.91 | -0.02 | 12.93 (12.69, 13.18) |
| **Calibration slope** | 1.00 (0.98, 1.03) | 0.0033 | 0.997 (0.995, 0.998) |
| **Calibration-in-the-large (intercept) (mmol/mol)** | -0.06 (-1.74, 1.62) | -0.2128 | 0.21 (0.18, 0.32) |

**b) External validation.** Model outcome is 6 month HbA1c.

| | CPRD validation cohort (n=9,376) | CANTATA D/D2 trials (n=1,755) | BI1245.20 trial (n=630) |
|---|---|---|---|
| **$R^2$** | 0.29 (0.27, 0.31) | 0.28 (0.24, 0.33) | 0.26 (0.20, 0.34) |
| **Root mean square error (mmol/mol)** | 13.2 | 9.1 | 8.5 |
| **Calibration slope** | 1.02 (0.99, 1.05) | 0.94 (0.88, 1.02) | 1.06 (0.93, 1.20) |
| **Calibration-in-the-large (intercept) (mmol/mol)** | -1.2 (-3.3, 0.9) | -1.0 (-5.1, 3.1) | -6.1 (-14.0, 1.8) |

**sFigure 3: Assessment of calibration between model predicted HbA1c 6 month outcome and observed HbA1c in derivation and validation cohorts**

**a) CPRD derivation cohort (n=14,069)**

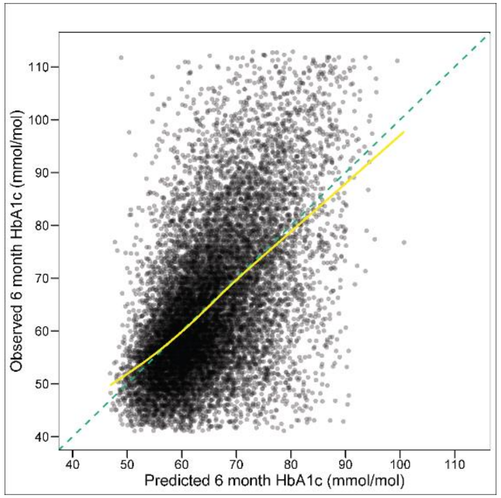


**b) CPRD validation cohort (n=9,376)**

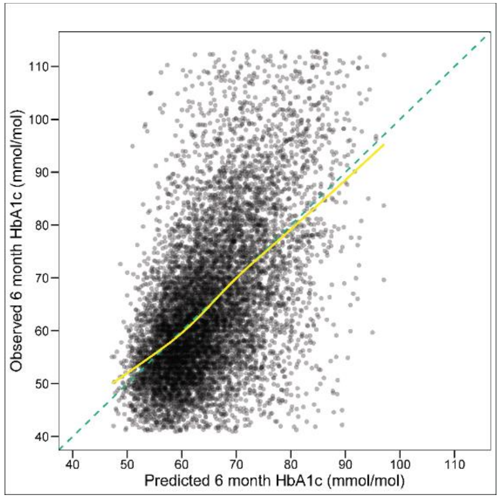


**c) CANTATA D/D2 trials (n=1,755)***

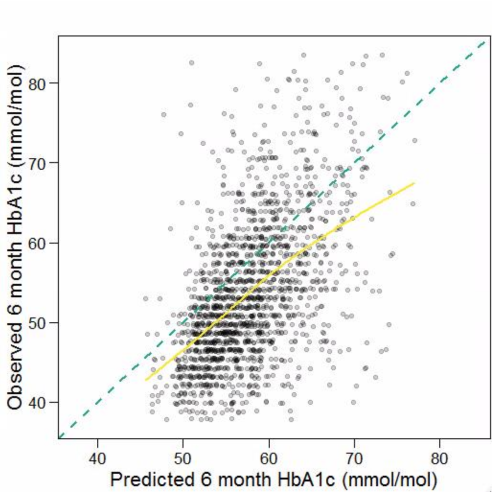


**d) BI1245.20 trial (n=630)***

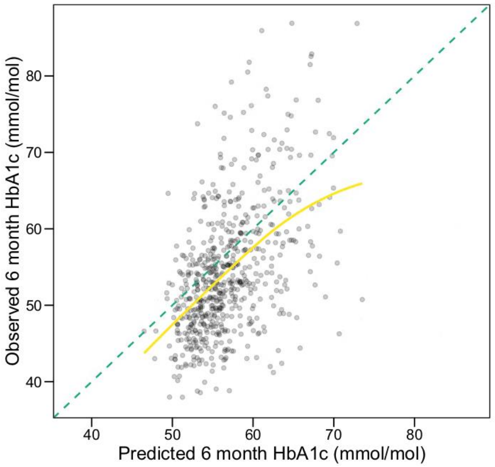


*In the trials, t

**sTable 5: Observed treatment effects across subgroups defined by clinical cut-offs of predicted treatment effects (data underlying Figure 4).** In CPRD, estimates are adjusted for clinical features in the treatment selection model (to improve precision and control for potential differences in covariate balance within subgroups). Trial data estimates are unadjusted**.**

#### a) CPRD validation sample (n=9,376).

| | Observed treatment difference (mmol/mol; negative favours SGLT2i) | | | | |
|---|---|---|---|---|---|
| **Predicted HbA1c difference** | **N patients** | **Treatment difference** | **Lower CI** | **Upper CI** | **p-value** |
| **Overall** | 9,376 | -4.7 | -5.4 | -4.2 | 0.000 |
| **Subgroup** | | | | | |
| SGLT2i benefit by any mmol/mol | 7,848 | -5.4 | -6.0 | -4.7 | <0.001 |
| SGLT2i benefit by ≥5 mmol/mol | 3,733 | -8.8 | -9.8 | -7.8 | <0.001 |
| SGLT2i benefit by 3-5 mmol/mol | 1,939 | -2.7 | -4.1 | -1.4 | <0.001 |
| SGLT2i benefit by 0-3 mmol/mol | 2,136 | -0.8 | -2.0 | 0.5 | 0.22 |
| DPP4i benefit by any mmol/mol | 1,528 | 0.5 | -1.5 | 2.4 | 0.62 |
| DPP4i benefit by 0-3 mmol/mol | 1,094 | 0.1 | -2.1 | 2.4 | 0.92 |
| DPP4i benefit by ≥3 mmol/mol | 434 | 2.0 | -2.3 | 6.3 | 0.36 |

#### b) CANTATA-D and CANTATA-D2 clinical trial participants (n=1,755).

| | Observed treatment difference (mmol/mol; negative favours SGLT2i) | | | | |
|---|---|---|---|---|---|
| **Predicted HbA1c difference** | **N patients** | **Treatment difference** | **Lower CI** | **Upper CI** | **p-value** |
| **Overall** | 1,755 | -1.4 | -2.2 | -0.6 | 0.003 |
| **Subgroup** | | | | | |
| SGLT2i benefit by any mmol/mol | 1,384 | -2.2 | -3.2 | -1.3 | <0.001 |
| SGLT2i benefit by ≥5 mmol/mol | 443 | -5.8 | -7.7 | -3.9 | <0.001 |
| SGLT2i benefit by 3-5 mmol/mol | 360 | -2.0 | -3.8 | -0.2 | 0.03 |
| SGLT2i benefit by 0-3 mmol/mol | 581 | 0.6 | -0.7 | 1.8 | 0.37 |
| DPP4i benefit by any mmol/mol | 371 | 1.4 | 0.1 | 2.6 | 0.03 |
| DPP4i benefit by 0-3 mmol/mol | 287 | 0.9 | -0.5 | 2.4 | 0.18 |
| DPP4i benefit by ≥3 mmol/mol | 84 | 2.3 | -0.8 | 5.3 | 0.14 |

#### c) BI1245.20 clinical trial participants (n=630).

| | Observed treatment difference (mmol/mol; negative favours SGLT2i) | | | | |
|---|---|---|---|---|---|
| **Predicted HbA1c difference** | **N patients** | **Treatment difference** | **Lower CI** | **Upper CI** | **p-value** |
| **Overall** | 630 | -1.1 | -2.6 | 0.5 | 0.19 |
| **Subgroup** | | | | | |
| SGLT2i benefit by any mmol/mol | 459 | -1.9 | -3.8 | 0.1 | 0.06 |
| SGLT2i benefit by ≥5 mmol/mol | 117 | -6.6 | -11.0 | -2.2 | 0.003 |
| SGLT2i benefit by 3-5 mmol/mol | 126 | -1.1 | -4.6 | 2.4 | 0.54 |
| SGLT2i benefit by 0-3 mmol/mol | 216 | 0.9 | -1.5 | 3.4 | 0.46 |
| DPP4i benefit by any mmol/mol | 171 | 1.5 | -0.7 | 3.7 | 0.17 |
| DPP4i benefit by 0-3 mmol/mol | 131 | 1.3 | -1.3 | 3.9 | 0.32 |
| DPP4i benefit by ≥3 mmol/mol | 40 | 2.6 | -1.6 | 6.8 | 0.22 |

**sFigure 4: Distribution of the predicted individualised treatment effect of SGLT2-inhibitor treatment compared to DPP4-inhibitor treatment in independent validation cohorts.** Negative values reflect a predicted glucose-lowering treatment benefit on SGLT2-inhibitor treatment, positive values reflect a predicted treatment benefit on DPP4-inhibitor treatment.

**a) CPRD validation cohort (n=9,376)**

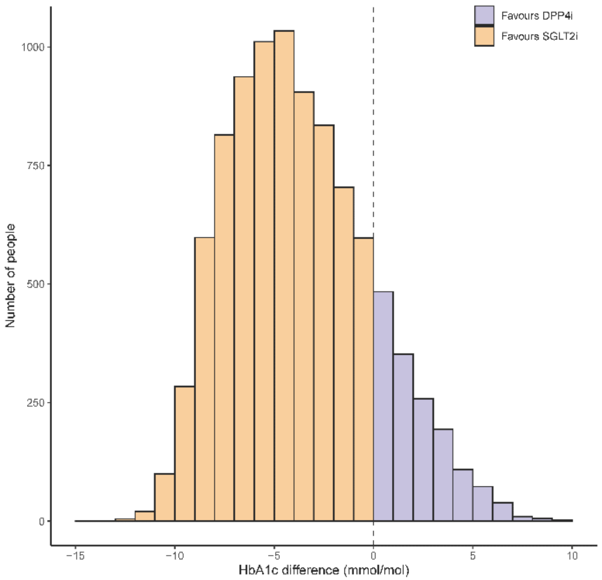


**b) CANTATA D/D2 trials (n=1,755)**

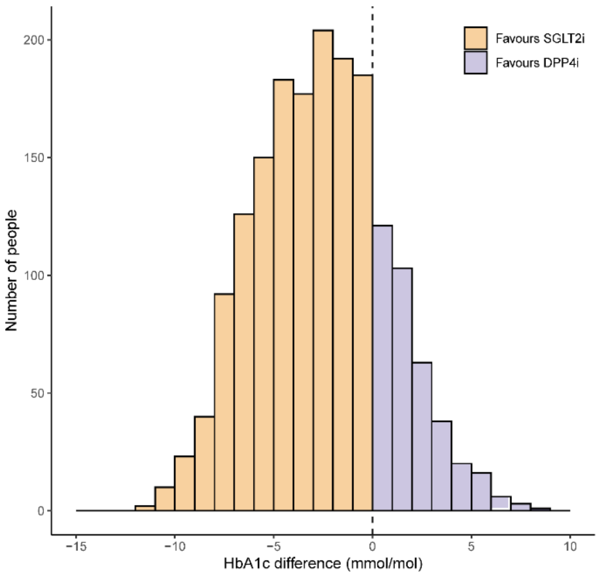


**c) BI1245.20 trial (n=630)**

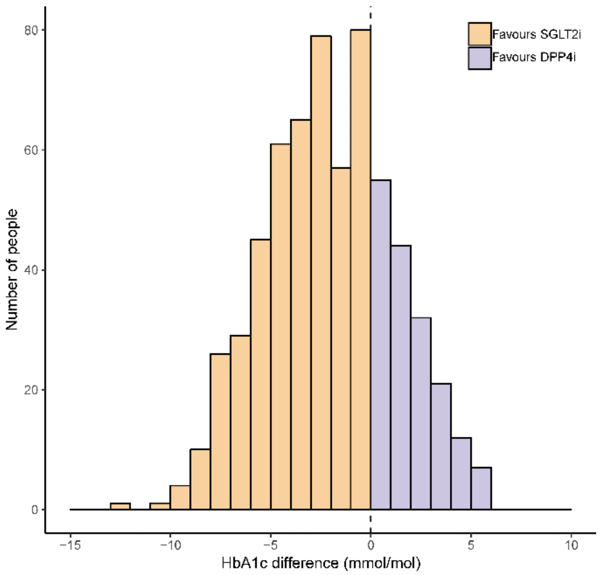

**sTable 6: Validation of 12 month HbA1c: Observed treatment effects across subgroups defined by clinical cut-offs of predicted treatment effects.** Estimates are unadjusted**.**

**a) CANTATA-D and CANTATA-D2 clinical trial participants (n=1,775).** 20 additional participants included who had an HbA1c outcome recorded post but not prior to 6 months.

| | Observed treatment difference (mmol/mol; negative favours SGLT2i) | | | | |
|---|---|---|---|---|---|
| **Predicted HbA1c difference** | **N patients** | **Treatment difference** | **Lower CI** | **Upper CI** | **p-value** |
| **Overall** | 1,775 | -2.4 | -3.4 | -1.4 | <0.001 |
| **Subgroup** | | | | | |
| SGLT2i benefit by any mmol/mol | 1,399 | -3.4 | -4.6 | -2.3 | <0.001 |
| SGLT2i benefit by ≥5 mmol/mol | 446 | -6.4 | -8.6 | -4.2 | <0.001 |
| SGLT2i benefit by 3-5 mmol/mol | 365 | -3.8 | -5.9 | -1.7 | <0.001 |
| SGLT2i benefit by 0-3 mmol/mol | 588 | 0.5 | -2.1 | 1.1 | 0.54 |
| DPP4i benefit by any mmol/mol | 376 | 0.7 | -0.8 | 2.2 | 0.34 |
| DPP4i benefit by 0-3 mmol/mol | 291 | 0.3 | -1.4 | 2.0 | 0.70 |
| DPP4i benefit by ≥3 mmol/mol | 85 | 1.8 | -1.3 | 4.8 | 0.25 |

**b) BI1245.20 clinical trial participants (n=630).**

| | Observed treatment difference (mmol/mol; negative favours SGLT2i) | | | | |
|---|---|---|---|---|---|
| **Predicted HbA1c difference** | **N patients** | **Treatment difference** | **Lower CI** | **Upper CI** | **p-value** |
| **Overall** | 630 | -2.1 | -3.7 | -0.4 | 0.01 |
| **Subgroup** | | | | | |
| SGLT2i benefit by any mmol/mol | 459 | -1.9 | -3.8 | 0.1 | 0.06 |
| SGLT2i benefit by ≥5 mmol/mol | 117 | -7.6 | -12.0 | -3.3 | <0.001 |
| SGLT2i benefit by 3-5 mmol/mol | 126 | -2.5 | -6.2 | 1.3 | 0.20 |
| SGLT2i benefit by 0-3 mmol/mol | 216 | -0.2 | -2.8 | 2.4 | 0.88 |
| DPP4i benefit by any mmol/mol | 171 | 0.9 | -1.8 | 3.5 | 0.52 |
| DPP4i benefit by 0-3 mmol/mol | 131 | 0.9 | -2.2 | 4.0 | 0.53 |
| DPP4i benefit by ≥3 mmol/mol | 40 | 0.7 | -4.7 | 6.0 | 0.80 |

**sTable 7: 6 month weight change and risk of treatment discontinuation, across subgroups defined by clinical cut-offs of predicted treatment benefit, in CPRD routine clinical data**

a) **Estimates of 6 month weight change.** Estimates include all patients with valid baseline data for glucose-lowering treatment selection model and with weight outcome recorded between 3 and 15 months after drug initiation, on unchanged glucose-lowering therapy. Estimates are adjusted for baseline weight, the number of currently prescribed glucose-lowering treatments, and the number of glucose-lowering drug classes ever prescribed.

| | Weight change kg (Median [IQR]) | | |
|---|---|---|---|
| **Predicted HbA1c difference** | **N patients** | **SGLT2** | **DPP4** |
| **Overall** | 15,627 | -3.7 (-4.3, -3.2) | -1.0 (-1.6, -0.5) |
| **Subgroup** | | | |
| SGLT2i benefit by any mmol/mol | 13,212 | -3.8 (-4.4, -3.3) | -1.0 (-1.6, -0.5) |
| SGLT2i benefit by ≥5 mmol/mol | 6,407 | -3.9 (-4.5, -3.4) | -1.0 (-1.6, -0.4) |
| SGLT2i benefit by 3-5 mmol/mol | 3,239 | -3.9 (-4.6, -3.4) | -1.0 (-1.6, -0.5) |
| SGLT2i benefit by 0-3 mmol/mol | 3,566 | -3.7 (-4.3, -3.2) | -1.1 (-1.6, -0.6) |
| DPP4i benefit by any mmol/mol | 2,415 | -3.0 (-3.5, -2.4) | -0.9 (-1.5, -0.5) |
| DPP4i benefit by 0-3 mmol/mol | 1,763 | -3.2 (-3.7, -2.7) | -1.0 (-1.5, -0.5) |
| DPP4i benefit by ≥3 mmol/mol | 652 | -2.4 (-3.0, -1.9) | -0.8 (-1.3, -0.4) |

b) **Estimates of treatment discontinuation within 6 months.** Estimates include all patients with valid baseline data for glucose-lowering treatment selection model and with 3 additional months of follow up to confirm treatment was truly discontinued. Estimates are adjusted for the number of currently prescribed glucose-lowering treatments, and the number of glucose-lowering drug classes ever prescribed.

| | Treatment discontinuation % (Median [IQR]) | | |
|---|---|---|---|
| **Predicted HbA1c difference** | **N patients** | **SGLT2** | **DPP4** |
| **Overall** | 28,514 | 16.1 (13.5, 20.3) | 14.4 (12.9, 16.7) |
| **Subgroup** | | | |
| SGLT2i benefit by any mmol/mol | 23,992 | 15.2 (13.2, 17.9) | 14.4 (12.9, 16.7) |
| SGLT2i benefit by ≥5 mmol/mol | 11,730 | 13.2 (12.1, 14.8) | 14.4 (13.0, 16.7) |
| SGLT2i benefit by 3-5 mmol/mol | 5,761 | 15.6 (14.4, 17.5) | 14.2 (12.8, 16.7) |
| SGLT2i benefit by 0-3 mmol/mol | 6,431 | 19.0 (17.3, 21.3) | 14.2 (12.8, 16.7) |
| DPP4i benefit by any mmol/mol | 4,592 | 26.8 (23.4, 31.0) | 14.8 (12.9, 16.8) |
| DPP4i benefit by 0-3 mmol/mol | 3,249 | 25.0 (22.5, 27.7) | 14.7 (12.8, 16.8) |
| DPP4i benefit by ≥3 mmol/mol | 1,343 | 33.1 (29.7, 36.9) | 14.9 (13.0, 16.9) |

**sFigure 5: Sensitivity analysis for 12 month weight change and risk of treatment discontinuation, across subgroups defined by clinical cut-offs of predicted treatment benefit, in CPRD routine clinical data**

a) **Weight change at 12 months (n=11,298).** Estimates include all patients with valid baseline data for glucose-lowering treatment selection model and with weight outcome recorded between 9 and 15 months after drug initiation, on unchanged glucose-lowering therapy. Estimates are adjusted for baseline weight, the number of currently prescribed glucose-lowering treatments, and the number of glucose-lowering drug classes ever prescribed.

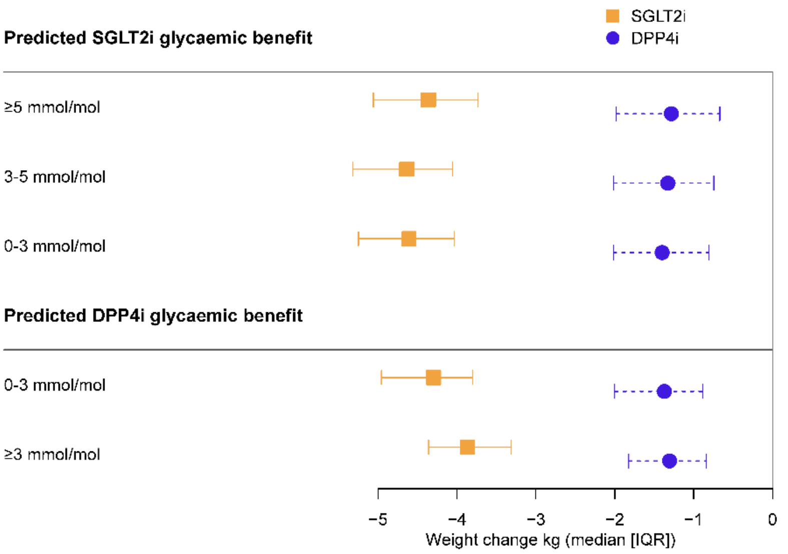


b) **Risk of treatment discontinuation within 12 months (n=23,739.** Estimates include all patients with valid baseline data for glucose-lowering treatment selection model and with 3 additional months of follow up to confirm treatment was truly discontinued. Estimates are adjusted for the number of currently prescribed glucose-lowering treatments, and the number of glucose-lowering drug classes ever prescribed.

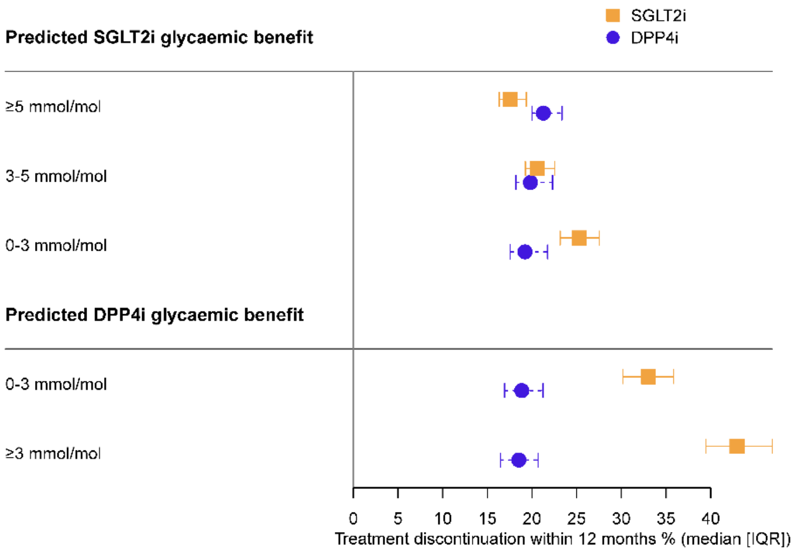

**sTable 8: Clinical characteristics of the entire CPRD primary care cohort by subgroups defined by model predicted HBA1c differences between therapies (n=36,454 with valid data to fit the treatment selection model).**

| | Predicted HbA1c benefit subgroup | | | | | | | | | |
|---|---|---|---|---|---|---|---|---|---|---|
| | SGLT2i ≥5 mmol/mol | | SGLT2i 3-5 mmol/mol | | SGLT2i 0-3 mmol/mol | | DPP4i 0-3 mmol/mol | | DPP4i ≥3 mmol/mol | |
| **n** | | **14860 (40.8%)** | | **7332 (20.1%)** | | **8245 (22.6%)** | | **4203 (11.5%)** | | **1814 (5.0%)** |
| **Clinical characteristics (median [IQR])** | | | | | | | | | | |
| Age (years) | | 55.0 [50.0, 61.0] | | 62.0 [57.0, 67.0] | | 68.0 [62.0, 73.0] | | 73.0 [69.0, 78.0] | | 79.0 [75.0, 84.0] |
| Duration of diabetes (years) | | 6.1 [3.4, 9.5] | | 7.6 [4.3, 11.3] | | 8.6 [5.1, 12.5] | | 9.9 [6.1, 14.0] | | 11.5 [7.5, 15.9] |
| Male sex (n [%]) | | 9892 (66.6) | | 4551 (62.1) | | 4827 (58.5) | | 2259 (53.7) | | 935 (51.5) |
| HbA1c (mmol/mol) | | 80.2 [72.0, 91.0] | | 70.0 [64.0, 79.0] | | 66.0 [61.0, 74.0] | | 64.0 [59.0, 70.0] | | 61.0 [57.0, 66.0] |
| BMI (kg/m2) | | 34.1 [30.7, 38.3] | | 32.1 [28.7, 36.6] | | 30.8 [27.7, 35.0] | | 29.2 [26.1, 33.2] | | 26.5 [24.0, 30.0] |
| eGFR (mL/min/1.3 m2) | | 97 [89, 104] | | 88 [77, 96] | | 79 [68, 89] | | 68 [59, 79] | | 59 [51, 68] |
| HDL-c (mmol/L) | | 1.0 [0.9, 1.2] | | 1.1 [0.9, 1.3] | | 1.1 [1.0, 1.3] | | 1.2 [1.0, 1.4] | | 1.3 [1.1, 1.5] |
| Triglycerides (mmol/L) | | 2.1 [1.5, 3.0] | | 1.9 [1.4, 2.7] | | 1.8 [1.3, 2.5] | | 1.7 [1.2, 2.3] | | 1.5 [1.1, 2.1] |
| ALT (IU/L) | | 37.0 [27.0, 52.0] | | 28.0 [21.0, 38.0] | | 24.0 [18.0, 33.0] | | 19.0 [15.0, 25.0] | | 15.0 [12.0, 20.0] |
| Albumin (g/L) | | 43.0 [40.0, 45.0] | | 42.0 [39.0, 45.0] | | 42.0 [39.0, 45.0] | | 42.0 [39.0, 45.0] | | 41.0 [38.0, 44.0] |
| Bilirubin (µmol/L) | | 9.0 [7.0, 12.0] | | 9.0 [6.0, 12.0] | | 9.0 [6.0, 12.0] | | 9.0 [6.0, 12.0] | | 8.8 [6.0, 11.0] |
| **Treatment received** | | | | | | | | | | |
| SGLT2-inhibitor | | 7399 (49.8%) | | 4173 (56.9%) | | 2888 (35.0% | | 925 (22.0%) | | 205 (11.3%) |
| DPP4-inhibitor | | 7461 (50.2%) | | 3159 (43.1%) | | 5357 (65.0%) | | 3278 (78.0%) | | 1609 (88.7%) |
| **Predicted 6 month outcomes by therapy** | | | | | | | | | | |
| Therapy | DPP4i | SGLT2i | DPP4i | SGLT2i | DPP4i | SGLT2i | DPP4i | SGLT2i | DPP4i | SGLT2i |
| HbA1c change (mmol/mol) | -8.5 [-13.2, -4.7] | -15.7[-20.8,-11.4] | -5.6 [-9.7, -2.2] | -9.7 [-13.8, -6.1] | -4.8 [-8.7, -1.6] | -6.5 [-10.4, -3.2] | -5.1 [-8.6, -1.8] | -3.8 [-7.4, -0.4] | -5.2 [-8.4, -1.6] | -0.4 [-4.0, 3.4] |
| Weight change (KG) | -0.9 [-1.5, -0.4] | -3.8 [-4.4, -3.3] | -1.0 [-1.6, -0.5] | -3.9 [-4.5, -3.4] | -1.0 [-1.6, -0.5] | -3.7 [-4.2, -3.2] | -0.9 [-1.5, -0.5] | -3.1 [-3.7, -2.6] | -0.8 [-1.3, -0.4] | -2.4 [-2.9, -1.9] |
| Treatment discontinuation (%) | 14.4 [13.0, 16.7] | 13.3 [12.1, 14.9] | 14.2 [12.8, 16.7] | 15.6 [14.4, 17.6] | 14.2 [12.8, 16.7] | 19.1 [17.3, 21.4] | 14.8 [12.8, 16.8] | 25.1 [22.6, 28.0] | 14.9 [13.0, 16.9] | 33.4 [29.8, 37.3] |
