## Supplementary Tables 2 for "Derivation and validation of a type 2 diabetes treatment selection algorithm for SGLT2-inhibitor and DPP4-inhibitor therapies based on glucose-lowering efficacy: cohort study using trial and routine clinical data"

| | Trial | Trial type | N SGLT2-inhibitor (final study cohorts) [1] | | | | | N DPP4-inhibitor (final study cohorts) [1] | | | | | | Background therapy | Primary outcome(s) | Study time period | Inclusion criteria[2] |
|---|---|---|---|---|---|---|---|---|---|---|---|---|---|---|---|---|---|
| | | | Canagliflozin | | Empagliflozin | | Any SGLT2-i | Sita-gliptin | Alogliptin | | | Lina-gliptin | Any DPP4-i | | | | |
| | | | 100mg | 300mg | 10mg | 25mg | | 100mg | 6.25mg | 12.5mg | 25mg | 5mg | | | | | |
| 1 | CANTATA-D (NCT01106677) | Efficacy, placebo-controlled and active comparator | 352 | 353 | | | 705 | 355 | | | | | 355 | Immediate-release metformin monotherapy (≥2,000 mg/day, or ≥1,500 mg/day if unable to tolerate higher dose).<br><br>(Participants on sulfonylurea in addition to metformin discontinued sulfonylurea.) | HbA1c change from baseline to 26 weeks | April 2010-August 2012 | • Aged 18-80<br>• HbA1c ≥7% and ≤10.5%<br>• Currently treated with metformin (maximum dose) alone or combined with sulfonylurea<br>**Exclusion:**<br>• Uncontrolled hyperglycaemia defined as repeated FPG ≥15.5mmol/L<br>• Uncontrolled hypertension<br>• eGFR <55mL/min (or <60 mL/min if based upon restriction in local label) or serum creatinine ≥124 µmol/L (men) or ≥115 µmol/L (women)<br>• Treatment with PPAR-γ agonist, insulin, another SGLT2-inhibitor or any other antihyperglycaemic agents except metformin alone or with sulfonylurea within 12 weeks<br>• Cardiovascular disease including MI, unstable angina, revascularisation procedure or cerebrovascular accident within 3 months<br>• Severe hypoglycaemia episode within 6 months |
| 2 | CANTATA-D2 (NCT01137812) | Efficacy, placebo-controlled and active comparator | | 359 | | | 359 | 356 | | | | | 356 | Combined metformin and sulfonylurea at maximum- or near-maximum doses (adjusted pre-run in if not already at these doses) | HbA1c change from baseline to 52 weeks | June 2010-March 2012 | • Aged 18+<br>• HbA1c ≥7% and ≤10.5%<br>• Currently treated with metformin alone or combined with sulfonylurea<br>**Exclusion:**<br>• Uncontrolled hyperglycaemia defined as repeated FPG ≥16.7mmol/L<br>• Uncontrolled hypertension<br>• eGFR <55mL/min (or <60 mL/min if based upon restriction in local label) or serum creatinine ≥124 µmol/L (men) or ≥115 µmol/L (women)<br>• Treatment with PPAR-γ agonist, insulin, another SGLT2-inhibitor or any other antihyperglycaemic agents except metformin and sulfonylurea within 12 weeks<br>• History of cardiovascular disease<br>• Severe hypoglycaemia episode within 6 months |
| 3 | EMPA-REG MONO (BI1245.20; NCT01177813) | Efficacy, placebo-controlled and active comparator | | | 214 | 291 | 505 | 219 | | | | | 219 | None (drug-naïve) | HbA1c change from baseline to 24 weeks | August 2010-March 2012 | • Aged 18+ (20+ in Japan only)<br>• HbA1c ≥7% and ≤10%<br>• BMI ≤45kg/m²<br>• Drug-naïve to antihyperglycaemic agents<br>**Exclusion:**<br>• Uncontrolled hyperglycaemia after overnight fast during placebo run-in<br>• eGFR <50mL/min at screening/placebo run-in<br>• Current treatment with systemic steroids or change in dosage of thyroid hormones within 6 weeks or any uncontrolled endocrine disorder except T2D<br>• Treatment with anti-obesity medication/other treatment leading to unstable body weight<br>• Any acute coronary syndrome], stroke, TIA within 3 months<br>• Bariatric/other GI surgeries within 2 years<br>• History of cancer<br>• Any disorders causing haemolysis or unstable red blood cells |
| 4 | CANTATA-SU (NCT00968812) | Efficacy, active comparator (sulfonylurea) | 470 | 469 | | | 939 | | | | | | | Immediate-release metformin monotherapy (≥2,000 mg/day, or ≥1,500 mg/day if unable to tolerate higher dose).<br><br>(Participants on combined metformin and a second antihyperglycaemic agent (not TZD as per inclusion criteria) discontinued second agent.) | HbA1c change from baseline to 52 weeks | August 2009-December 2011 (core period) or January 2013 (52-week extension) | • Aged 18-80<br>• HbA1c ≥7% and ≤9.5%<br>• BMI ≥22 and ≤45kg/m²<br>• Currently treated with metformin (maximum dose) alone or combined with another antihyperglycaemic agent<br>**Exclusion:**<br>• Uncontrolled hyperglycaemia defined as repeated FPG ≥15mmol/L<br>• History of active proliferative diabetic retinopathy<br>• eGFR <55mL/min (or <60 mL/min if based upon restriction in local label) or serum creatinine ≥124 µmol/L (men) or ≥115 µmol/L (women)<br>• Treatment with TZD within 16 weeks<br>• Hereditary glucose-galactose malabsorption or primary renal glucosuria<br>• Renal disease requiring immunosuppressive therapy within 12 months or a history of dialysis or renal transplant<br>• Severe hypoglycaemia episode within 6 months |
| 5 | CANTATA-M (NCT01081834) | Efficacy, placebo-controlled | 229 | 230 | | | 459 | | | | | | | None (participants on antihyperglycaemic agents as per inclusion criteria discontinued these) | HbA1c change from baseline to 26 weeks | March 2010-March 2012 | • Aged 18-80<br>• Main study: HbA1c ≥7% and ≤10% on no antihyperglycaemic agent/HbA1c ≥6.5% and ≤9.5% on antihyperglycaemic agent monotherapy or combined low dose metformin and sulfonylurea; FPG ≤15mmol/L<br>• High Glycaemic Substudy: HbA1c >10% and ≤12% and FPG ≤19.4mmol/L<br>**Exclusion:**<br>• eGFR <50mL/min<br>• Treatment with PPAR-γ agonist, insulin, another SGLT2-inhibitor or any other antihyperglycaemic agents except as specified above within 12 weeks<br>• Cardiovascular disease including MI, unstable angina, revascularisation procedure or cerebrovascular accident within 3 months<br>• Hereditary glucose-galactose malabsorption or primary renal glucosuria<br>• Severe hypoglycaemia episode within 6 months |
| 6 | Efficacy, Safety, and Tolerability of Canagliflozin vs. Placebo in the Treatment of Older Subjects… (NCT01106651) | Efficacy, placebo-controlled | 236 | 228 | | | 303 | | | | | | | Any (usual care) | HbA1c change from baseline to 26 weeks | April 2010-November 2011 (core period) or June 2013 (78-week extension) | • Aged 55-80 (women at least 3 years postmenopausal)<br>• HbA1c ≥7% and ≤10%<br>• BMI ≥20 and ≤40kg/m²<br>• Fasting fingerstick blood glucose ≥6.1mmol/L<br>**Exclusion:**<br>• Uncontrolled hyperglycaemia defined as repeated FPG ≥15mmol/L<br>• Uncontrolled hypertension<br>• eGFR <50mL/min<br>• On metformin and serum creatinine ≥124 µmol/L (men) or ≥115 µmol/L (women)<br>• Cardiovascular disease including MI, unstable angina, revascularisation procedure or cerebrovascular accident within 3 months<br>• History of New York Heart Association Class III–IV cardiac disease<br>• Severe hypoglycaemia episode within 6 months |
| 7 | CANTATA-MSU (NCT01106625) | Efficacy, placebo-controlled | 149 | 154 | | | 303 | | | | | | | Combined metformin and sulfonylurea at maximum- or near-maximum doses (adjusted pre-run in if not already at these doses) | HbA1c change from baseline to 26 weeks | April 2010-April 2012 | • Aged 18-80<br>• HbA1c ≥7% and ≤10.5%<br>• Currently treated with combined metformin and sulfonylurea<br>**Exclusion:**<br>• Uncontrolled hyperglycaemia defined as repeated FPG ≥15mmol/L<br>• Uncontrolled hypertension<br>• eGFR <55mL/min (or <60 mL/min if based upon restriction in local label) or serum creatinine ≥124 µmol/L (men) or ≥115 µmol/L (women)<br>• Treatment with any other antihyperglycaemic agents except metformin and sulfonylurea within 12 weeks<br>• Severe hypoglycaemia episode within 6 months |
| 8 | EMPA-REG PIO (BI1245.19; NCT01210001) | Efficacy, placebo-controlled | | | 159 | 162 | 321 | | | | | | | Pioglitazone alone or in combination with metformin | HbA1c change from baseline to 24 weeks (for any background therapy and for pioglitazone + metformin subset) | September 2010-April 2012 | • Aged 18+<br>• BI1245.19: HbA1c ≥7% and ≤10%; BI1245.23: ≤11% for randomised arm, >11% for open-label arm<br>• BMI ≥45kg/m²<br>• Current treatment matches trial background therapy<br>**Exclusion:**<br>• Uncontrolled hyperglycaemia after overnight fast during placebo run-in<br>• eGFR <30mL/min at screening/placebo run-in<br>• Current treatment with systemic steroids or change in dosage of thyroid hormones within 6 weeks or any uncontrolled endocrine disorder except T2D<br>• Treatment with anti-obesity medication/other treatment leading to unstable body weight<br>• MI, stroke, TIA within 3 months<br>• Bariatric/other GI surgeries within 2 years<br>• History of cancer<br>• Any disorders causing haemolysis or unstable red blood cells |
| 9 | EMPA-REG METSU (BI1245.23; NCT01159600) | Efficacy, placebo-controlled | | | 434 | 581 | 1,015 | | | | | | | Immediate-release metformin (≥1500mg/day or maximum dose) alone or in combination with sulfonylurea (≥half maximum dose) | HbA1c change from baseline to 24 weeks | July 2010-February 2012 | |
| 10 | EMPA-REG OUTCOME (BI1245.25; NCT01131676) | CV outcomes, placebo-controlled | | | 1,100 | 1,110 | 2,210 | | | | | | | Any (usual care) | Time to first occurrence of 3-point MACE (composite of CV death, non-fatal MI, and non-fatal stroke) | July 2010-April 2015 | • Aged 18+<br>• HbA1c ≥7% and ≤10%, or ≥7% and ≤9% if drug-naïve<br>• BMI ≤45kg/m²<br>• High cardiovascular risk defined as ≥1 of the following (all >2 months prior): history of MI; evidence of single-vessel or multi-vessel coronary artery disease; unstable angina with evidence of single- or multi-vessel coronary artery disease; history of stroke (ischemic or hemorrhagic); occlusive peripheral artery disease<br>**Exclusion:**<br>• Uncontrolled hyperglycaemia after overnight fast during placebo run-in<br>• eGFR <30mL/min at screening/placebo run-in<br>• Planned cardiac surgery or angioplasty within 3 months<br>• Current treatment with systemic steroids or change in dosage of thyroid hormones within 6 weeks or any uncontrolled endocrine disorder except T2D<br>• Treatment with anti-obesity medication/other treatment leading to unstable body weight<br>• Acute coronary syndrome, stroke, TIA within 2 months<br>• Bariatric/other GI surgeries within 2 years<br>• History of cancer<br>• Any disorders causing haemolysis or unstable red blood cells |
| 11 | Linagliptin in Combination With Metformin and a Sulfonylurea… (BI1218.18; NCT00602472) | Efficacy, placebo-controlled | | | | | | | | | | 774 | 774 | Combined metformin (≥1500mg/day or maximum dose) and sulfonylurea (maximum dose) | HbA1c change from baseline to 24 weeks | February 2008-May 2009 | • Aged 18-80<br>• HbA1c ≥7% and ≤10%<br>• BMI ≤40kg/m²<br>• Current treatment matches trial background therapy<br>**Exclusion:**<br>• Renal failure, or renal impairment defined as serum creatinine ≥1.5 mg/dL<br>• Treatment with TZD, GLP-1 analogue, or insulin within 3 months<br>• Treatment with anti-obesity drugs (sibutramine, rimonabant, orlistat) within 3 months<br>• Current treatment with systemic steroids or change in dosage of thyroid hormones within 6 weeks<br>• MI, stroke, TIA within 6 months<br>• Dehydration<br>• Current acute or chronic metabolic acidosis |
| 12 | Efficacy and Safety of Linagliptin vs. Placebo… (BI1218.16; NCT00621140) | Efficacy, placebo-controlled | | | | | | | | | | 326 | 326 | None (participants on antihyperglycaemic agents as per inclusion criteria discontinued these) | HbA1c change from baseline to 24 weeks | February 2008-May 2009 | • Aged 18-80<br>• HbA1c ≥6.5% and ≤9%, or ≥7% and ≤10% if treatment-naïve<br>**Exclusion:**<br>• Treatment with >1 oral antidiabetic agent within 10 weeks<br>• Treatment with TZD, GLP-1 analogue, or insulin within 3 months<br>• Treatment with anti-obesity drugs (sibutramine, rimonabant, orlistat) within 3 months<br>• Current treatment with systemic steroids or change in dosage of thyroid hormones within 6 weeks<br>• MI, stroke, TIA within 6 months |
| 13 | Efficacy and Safety of Linagliptin vs. Placebo Added to Metformin Background Therapy… (BI1218.17; NCT00601250) | Efficacy, placebo-controlled | | | | | | | | | | 506 | 506 | Metformin monotherapy (≥1500mg/day or maximum dose).<br><br>(Participants on an additional oral antidiabetes agents as per inclusion criteria discontinued these.) | HbA1c change from baseline to 24 weeks | January 2008-May 2009 | • Aged 18-80<br>• HbA1c ≥6.5% and ≤9% if undergoing wash-out of previous medication, or ≥7% and ≤10% otherwise<br>• BMI ≤40kg/m²<br>• Currently treated with metformin (maximum dose) alone or combined with another oral antidiabetes agent<br>**Exclusion:**<br>• Renal failure or renal impairment<br>• Treatment with TZD, injectable GLP-1 analogue, or insulin within 3 months<br>• Treatment with anti-obesity drugs (sibutramine, rimonabant, orlistat) within 3 months<br>• Current treatment with systemic steroids or change in dosage of thyroid hormones within 6 weeks<br>• MI, stroke, TIA within 6 months<br>• Unstable or acute congestive heart failure<br>• Dehydration<br>• History of acute or chronic metabolic acidosis<br>• Hereditary galactose intolerance |
| 14 | Efficacy and Safety of Linagliptin in Combination With Metformin (BI1218.20; NCT00622284) | Efficacy, placebo-controlled and active comparator (sulfonylurea) | | | | | | | | | | 759 | 759 | Metformin monotherapy (≥1500mg/day or maximum dose).<br><br>(Participants on an additional oral antidiabetes agents as per inclusion criteria discontinued these.) | HbA1c change from baseline to 52 weeks | February 2008-December 2010 | • Aged 18-80<br>• HbA1c ≥6% and ≤9% if treated with metformin and another oral antidiabetes agent,, or ≥6.5% and ≤10% if treated with metformin alone<br>• BMI ≤40kg/m²<br>• Currently treated with metformin (maximum dose) alone or combined with another oral antidiabetes agent<br>**Exclusion:**<br>• Renal failure or renal impairment<br>• Treatment with TZD, GLP-1 analogue, or insulin within 3 months<br>• MI, stroke, TIA within 6 months |

BMI, body mass index; DPP4-i(nhibitor), dipeptidyl-peptidase 4 inhibitor; eGFR, estimated glomerular filtration rate; FPG, fasting plasma glucose; GLP-1, glucagon-like peptide-1; HbA1c, glycated haemoglobin, type A1c; MACE, major adverse cardiovascular event; MI, myocardial infarction; PPAR-γ, peroxisome proliferator-activated receptor gamma ; SGLT2-i(nhibitor), sodium-glucose cotransporter-2 inhibitor; T2D, Type 2 diabetes; TIA, transient ischaemic attack; TZD, thiazolidinedione.

[1]Ns are participants included in the mixed effects models and subsequent meta-analysis. This includes participants who were not insulin-treated and with at least one on-treatment HbA1c between randomisation and 6 months (trials 3,8,9: HbA1c measurements excluded after changes in study medication (including dose changes); trial 10: HbA1c measurements excluded after changes in study medication (including dose changes) or changes in background medication (not including dose changes). Trials 1,2,4-7: treatment group is as randomised; trials 3,8-14: treatment group is actual study medication given.

[2]All trials:
• Diagnosis of Type 2 diabetes mellitus
• Background therapy as stated in table and unchanged for up to 12 weeks prior to randomisation (varies between trials)
**Exclusion:**
• (Symptoms of) Type 1 diabetes mellitus or other type of diabetes mellitus
• Contraindications (according to local market) to assigned background or study medications
• Liver disease defined by elevated ALT, AST and/or alkaline phosphatase
• Nursing or pregnant women; or premenopausal women of child bearing potential either intending to become pregnant during the trial period and/or not practicing an acceptable method of birth control
• History of alcohol or drug abuse
• Participation in another trial with an investigational drug within preceding 30 days
• Any other clinical condition that would jeopardise patient safety while participating in this clinical trial
